# Pseudo-*p*-Value-Based Clumping Enhanced Proteome-wide Mendelian Randomization with Application in Identifying Coronary Heart Disease-Associated Plasma Proteins

**DOI:** 10.1101/2025.01.13.25320450

**Authors:** Yuquan Wang, Yunlong Cao, Dong Chen, Dapeng Shi, Liwan Fu, Anqi Chen, Siyuan Shen, Yue-Qing Hu

## Abstract

Mendelian randomization (MR) is a powerful tool for causal inference in epidemiology. However, the presence of weak instrumental variables (IVs) and pleiotropy can lead to biased causal effect estimates. To address these issues, we develop MR-GMM, a novel MR method based on a Gaussian Mixture Model. MR-GMM classifies IVs into four categories—invalid, valid, invalid&null, and null IVs— and models their effects using a two-dimensional spike-and-slab distribution. Simulation studies demonstrate the high efficiency and robustness of MR-GMM compared to existing methods. More importantly, we propose a pseudo-***p***-value-based linkage disequilibrium (LD) clumping procedure to address selection bias. This refined procedure is capable of enhancing the performance of MR-GMM as well as many existing MR methods in real-world scenarios. Applying MR-GMM in a large-scale proteome-wide MR study, we identify 45 coronary heart disease-associated plasma proteins. Subsequent network and enrichment analyses highlight the potential of these proteins as biomarkers for disease diagnosis and therapeutic development.

## 1 Introduction

Genome-wide association studies (GWASs) have achieved enormous success in identifying genetic risk factors for complex traits, advancing our understanding of disease mechanisms and promoting drug development [1]. By employing genetic variants as instrumental variables (IVs), Mendelian randomization (MR) reuses GWAS summary statistics to infer possible causal relationships between exposures and outcomes [2]. As summary statistics from GWAS are more readily available compared to individual-level data, two-sample MR (2SMR) has recently gained widespread applications in the field of epidemiology [3].

For robust causal inference, IVs in MR analyses must satisfy three fundamental criteria (Figure 1a): (i) it must be associated with the exposure (Relevance); (ii) it must be independent of all unmeasured confounders of exposure and outcome (Random assignment); (iii) it must affect the outcome sorely through the exposure, not through any other pathway (Exclusion restriction) [4, 5]. Single nucleotide polymorphisms (SNPs) are widely used as IVs in MR studies because GWASs have shown their association with various exposures, and they are randomly inherited from parental chromosomes, which supports their alignment with these assumptions [6, 7].

**Fig 1:**
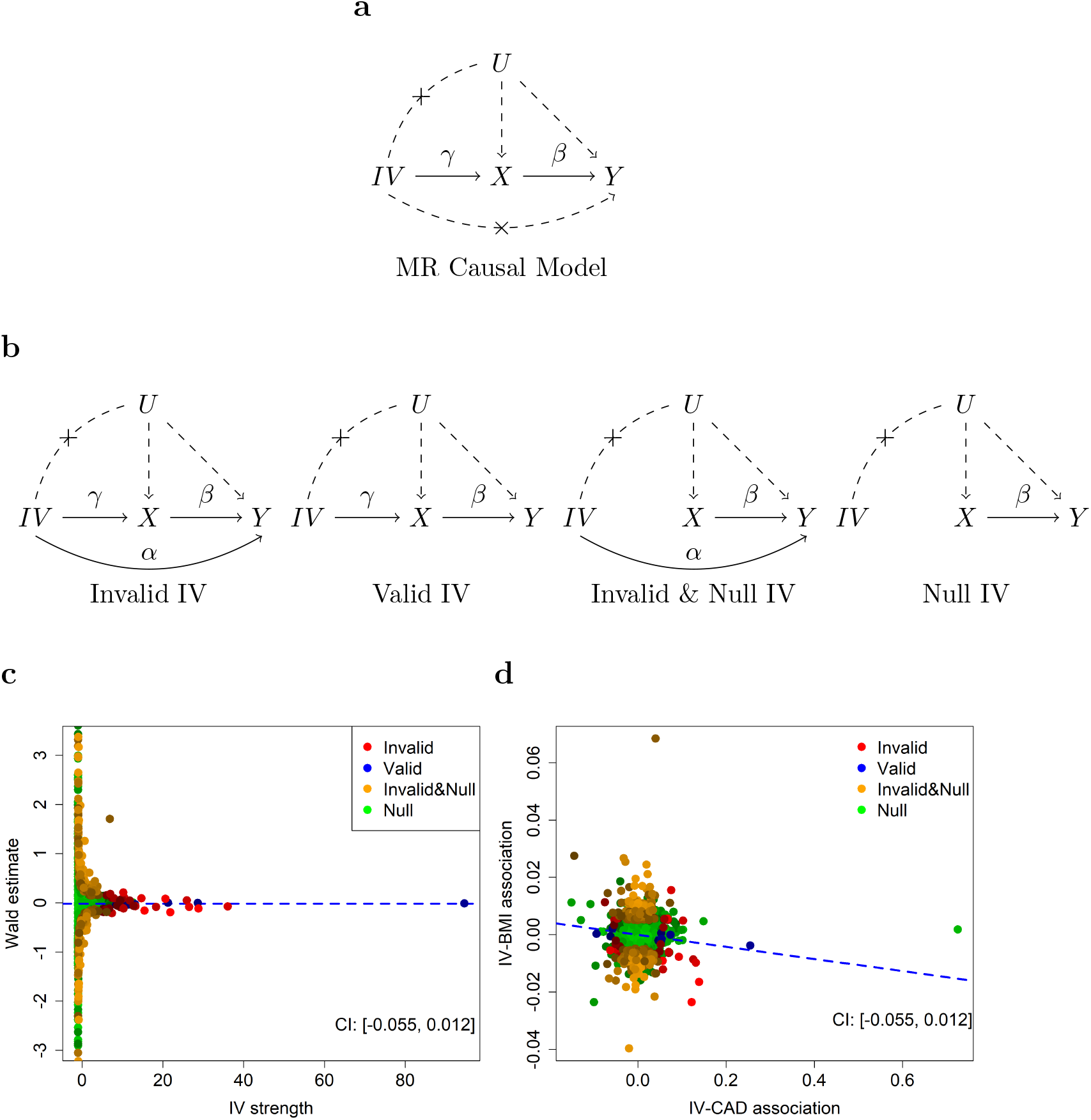
The causal models of valid IVs and MR-GMM are illustrated using the *cad*.*bmi* dataset from the *mr*.*raps* package. This dataset demonstrates a reverse causal relationship, where coronary artery disease (CAD) serves as the exposure, and body mass index (BMI) serves as the outcome. **a**. The three criteria for valid IVs in traditional MR causal models. **b**. The causal models of the four components in MR-GMM. The invalid IV is associated with the exposure but have a direct effect on the outcome, the valid IV satisfies the three criteria, the invalid & null IV only has a direct effect on the outcome, and the null IVs is neither correlated with the exposure nor the outcome. **c**. The Wald estimates for individual SNPs are plotted against their corresponding IV strengths in the *cad*.*bmi* dataset. The distribution in the plot demonstrates that the causal estimate obtained by MR-GMM is primarily influenced by the valid IVs. **d**. The scatter plot of IV3-0BMI associations against IV-CAD associations reveals distinct patterns: invalid & null IVs are vertically distributed, reflecting their direct effect on the outcome but no association with the exposure, while null IVs are clustered near the origin in a circle, indicating no association with either the exposure or the outcome. The 95% confidence interval for the causal effect estimated by MR-GMM includes the true value of *β* = 0, indicating that the method successfully captures the true causal effect.

In practice, these assumptions are often suspicious. Due to the polygenic nature of complex traits, the average effect size of SNPs tends to be small [5]. Consequently, increasing the number of SNPs is sometimes necessary to enhance the statistical power of MR analyses [8, 9], which can, however, introduce weak or pleiotropic SNPs, potentially compromising validity. Furthermore, the sample sizes of GWASs used in MR analyses largely influence the reliability of the results. Summary statistics derived from smaller sample sizes typically contain substantial measurement errors, which reduce the strength of IVs and consequently lower statistical power. Classic methods like MR-Egger and weighted median estimator were developed to mitigate pleiotropic effects. However, they failed to account for measurement errors in GWAS summary statistics [10, 11], yielding biased causal effect estimates that are often attenuated towards zero [5, 12]. Debiased Inverse-Variance Weighted (DIVW) estimator improves upon IVW by simultaneously accounting for measurement errors and systematic pleiotropy, offering more reliable causal inference [12]. The Robust Adjusted Profile Score (RAPS) method tackled idiosyncratic pleiotropy by applying robust loss functions [5]. The authors also highlighted that linkage disequilibrium (LD) clumping in MR analyses could introduce selection bias, sometimes referred to as the “winner’s curse”, which further biases causal effect estimates towards zero [5]. The Constrained Maximum Likelihood-based (cML) method is capable of handling correlated pleiotropy by modeling pleiotropy as fixed effects and leveraging the plurality rule [13]. MR-APSS method further corrects for the selection bias by applying a truncated normal mixture distribution, providing a more robust estimate of causal effects [14].

In this paper, we develop a MR method based on a Gaussian Mixture Model, denoted MR-GMM. By employing a two-dimensional spike-and-slab distribution to model both the SNP-exposure associations and SNP-outcome pleiotropy (Figure 1b), MR-GMM enables robust causal effect estimation even in the presence of many weak IVs, while maintaining high statistical efficiency. Unlike many existing mixture models [15, 16], MR-GMM further incorporates a Dirichlet prior to penalize the likelihood function of the GMM, ensuring component probabilities remain bounded away from 0 and 1 and thus enhancing the stability of statistical inference. More importantly, to address selection bias, we propose a novel pseudo-*p*-value-based LD clumping procedure inspired by RIVW [17]. This procedure avoids the selection bias introduced by the use of *p*-value-based LD clumping while also reducing measurement errors. We demonstrate the high efficiency and robustness of MR-GMM through simulation studies that included scenarios with weak IVs and pleiotropy. To validate the proposed LD clumping procedure, we use MR-GMM to estimate identity causal effects under varying *p*-value thresholds, demonstrating its advantages over traditional *p*-value-based clumping procedure. Additionally, we assess the sensitivity of MR-GMM by testing causal relationships between 81 different exposures and a negative control outcome. Finally, in a large-scale proteome-wide MR analysis, MR-GMM identified 45 coronary heart disease (CHD)-associated plasma proteins out of 2992 candidates, providing additional biomarkers for the diagnosis and prediction of CHD, as well as new insights for the development of targeted drugs.

## 2 Results

### 2.1 An overview of MR-GMM method

We classify IVs into four distinct classes to account for all possible behaviors of SNPs: (i) invalid IVs, which are associated with the exposure but have a direct effect on the outcome; (ii) valid IVs, which satisfy the three criteria; (iii) invalid & null IVs, which only have a direct effect on the outcome; (iv) null IVs, which are uncorrelated with both the exposure and the outcome (Figure 1b). Simulation studies and practical applications will show that this detailed classification of SNPs is able to enhance the power of MR analyses. The InSIDE (INstrument Strength Independent of Direct Effect) assumption is invoked here for model identifiability, which corresponds to the independence of IVs and unmeasured confounders in Figure 1b [11].

We use a two dimensional spike-and-slab distribution to model the four classes. Specifically, we assume the relationship between Γ_*j*_ and *γ*_*j*_—the effect of the *j*th IV on the outcome and the exposure, respectively—as follows:

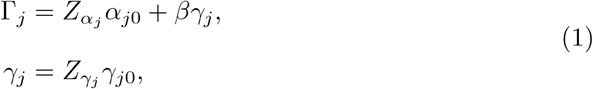

where *β* is the causal effect of interest, *γ*_*j*0_, *α*_*j*0_ are latent effects corresponding to IV-exposure association and IV-outcome pleiotropy, respectively. The binary indicators *Z*_*γj*_ = 1, 0 and *Z*_*αj*_ = 1, 0 specify whether the *j*th IV is relevant/invalid (slab) or not (spike). With different combinations of indicators *Z*_*γj*_ and *Z*_*αj*_, the four IV classes can be fully delineated. The latent effects are assumed to independently follow a specified distribution (usually normal distribution). We employ a spike-and-slab distribution instead of a fixed-effects model because the former is better suited for handling null and weak SNPs, which are common in GWAS summary statistics. Fixed-effects models typically rely on a sparsity assumption for pleiotropy and manage it through penalties on pleiotropic effects [13]. These penalties often require an additional beta-min assumption, which stipulates that the minimum effect size must exceed a threshold determined by sample size [18]. This assumption can be restrictive and less applicable to scenarios with weak or highly variable effects.

We illustrate the four IV classes using a real data example from the *mr*.*raps* package [5], focusing on the reverse causation of coronary artery disease (CAD) on body mass index (BMI). MR-GMM provides an interval estimate [−0.055, 0.012] for causal effect, including 0 as expected. Figures 1c and 1d depict the 1625 SNPs from *cad*.*bmi* dataset, showing their influence on MR-GMM in different aspects. It is worth noting that MR-GMM does not truly cluster SNPs. Rather, it provides the posterior probability of each SNP belonging to a specific class. In the plots, the class of each SNP corresponds to the one with the highest posterior probability. The brighter the color, the higher the probability that the SNP belongs to the indicated class. Both plots reveal that only a small proportion of the 1625 SNPs qualify as valid IVs by MR-GMM, with the majority classified as null or invalid & null IVs. In Figure 1c, the point estimate of MR-GMM aligns closely with Wald ratio estimates derived from the strongest IVs. Whereas Figure 1d demonstrates that MR-GMM primarily relies on SNPs identified as valid IVs, showing less influence from other IV classes. These plots show the necessity of accounting for the other three classes of IVs in MR analyses.

### 2.2 Assessing the statistical performance of MR-GMM through simulation studies

We designed four simulation scenarios to compare the statistical performance of MR-GMM with six existing methods: DIVW [12], MR-Egger [10], WME (weighted median estimator) [11], APS (RAPS method accounts for systematic pleiotropy only) [5], cML-MA-BIC [13], and MR-APSS (LD scores set to 1) [14]. The four scenarios—Balanced, Valid, Invalid & Null, and Null—correspond to GWAS summary data with equal proportions of the four classes IVs, the highest proportion of valid IVs, the highest proportion of invalid and null IVs, and the highest proportion of null IVs, respectively. The number of IVs was set to *m* = 500. The IV strength was calculated using the definition of average strength of IVs, *κ*, provided by RAPS [5]:

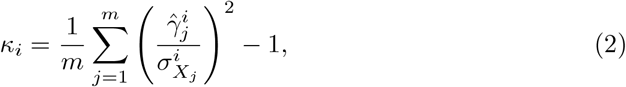

and taking mean across 1000 replications 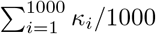, where 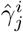 is the observed effect size of *j*th SNP on exposure in *i*th replication and 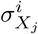 is the standard error of 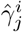. As expected, the IV strength was highest in the Valid scenario and lowest in the Invalid & Null and Null scenarios (Figure 2).

**Fig 2:**
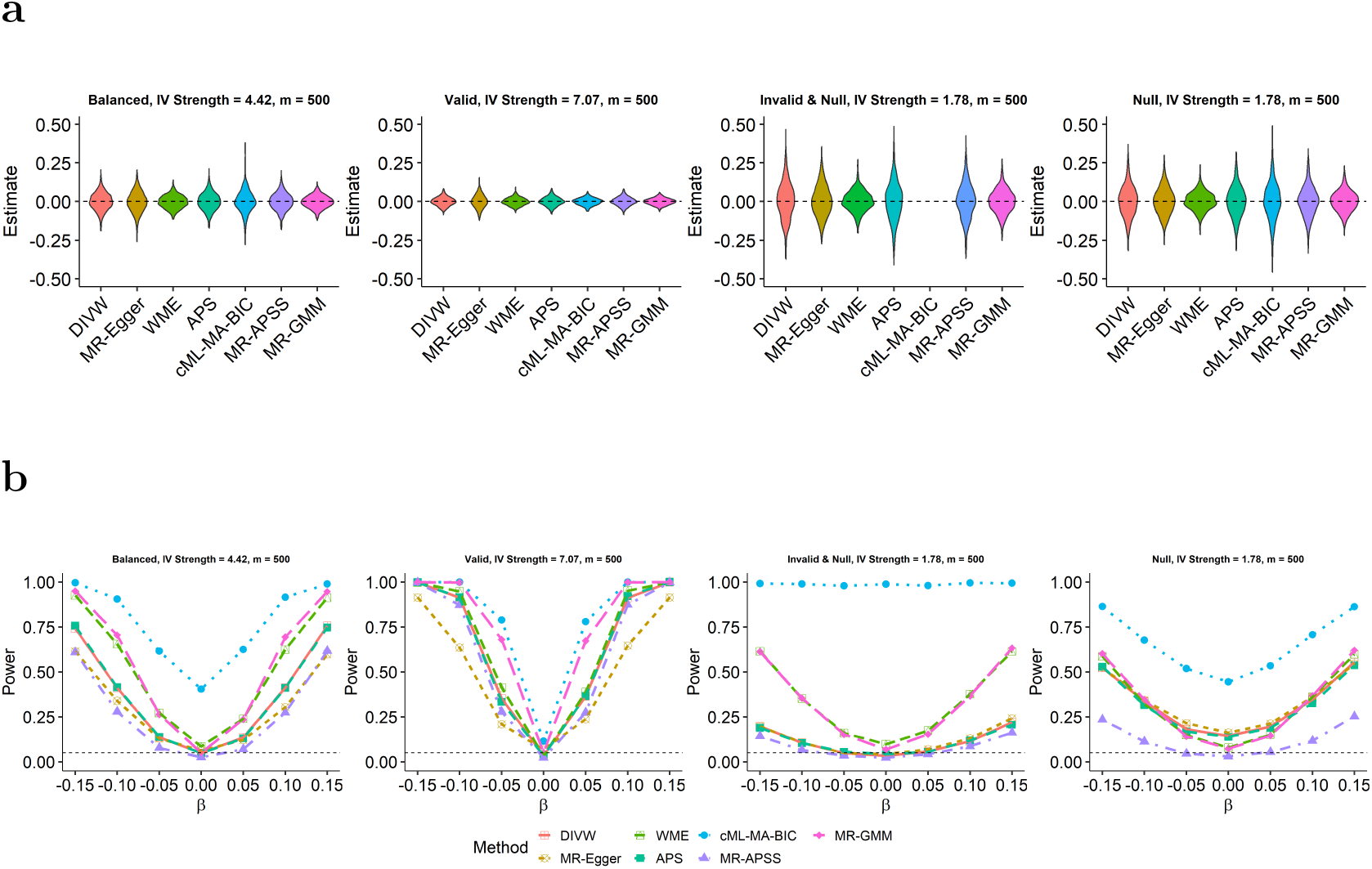
Benchmark existing MR methods in various simulation scenarios. Violin plots depict the point estimates from 1000 replications when the true causal effect *β* = 0, indicated by the dashed line. The violin plot for cML-MA-BIC is absent under the Invalid & Null scenario due to its poor performance in this scenario. **b**. The power of compared methods, calculated as the proportion of simulations in which the null hypothesis *H*_0_ : *β* = 0 is rejected across 1000 replications.

We first compared the point estimates of all methods across the four scenarios using violin plots shown in Figure 2a. The true parameter value, *β* = 0, is indicated by the horizontal dashed line. Among the methods, MR-GMM consistently displayed the highest density at the true value, highlighting its minimal bias and smallest standard error (SE) relative to the others. WME outperformed DIVW, MR-Egger, APS, and MR-APSS, though its density was asymmetrically distributed around the true value due to its basis on a median estimator. cML-MA-BIC exhibited the poorest performance, likely due to its basis on a fixed-effects model, which is less capable of addressing weak pleiotropy [13]. Moreover, the comparison across scenarios reveals that the SEs of causal effect estimates decrease as the average strength of the IVs increases. This observation underscores the importance of selecting as many strong IVs as possible in Mendelian randomization (MR) studies to improve the precision. Similar results for point estimates were observed under other values of *β*, as presented in Figure S1.

In terms of type I error rates, corresponding to the power at *β* = 0 in Figure 2b, MR-GMM and MR-APSS demonstrated consistent control of type I error across all scenarios. WME slightly exceeded the 0.05 threshold in the Invalid & Null scenario, while DIVW, MR-Egger, and APS showed marginally higher error rates than 0.05 in the Null scenario. In contrast, cML-MA-BIC exhibited the highest type I error among all methods. It approached the 0.05 threshold only in the Valid scenario but was significantly inflated in other cases. Next, we evaluated the power of all methods. Apart from cML-MA-BIC, which failed to control type I error in most scenarios, MR-GMM demonstrates the highest power. It is followed by DIVW, WME, and APS, while MR-Egger and MR-APSS were the most conservative among all methods. MR-APSS outperforms MR-Egger only in the Valid scenario. These simulation studies highlight that MR-GMM produces distributions with minimal variation and the highest density around the true parameter, reflecting reduced bias, lower variance, and superior power compared to other methods.

### 2.3 Pseudo-*p*-value-based LD clumping versus *p*-value-based LD clumping: illustrated through the estimation of identity causal effects

Generally, selection bias in *p*-value-based LD clumping arises from two main sources. One is the *p*-value threshold for selecting strong IVs, and the other lies in the selection of the index SNP within a genomic region. To illustrate the two-part selection bias introduced by *p*-value-based LD clumping procedure, we assessed the performance of all methods in estimating identity causal effects of BMI on BMI and HDL (high-density lipoprotein) on HDL under different IV selection thresholds.

As shown in Figure 3a, when the pseudo-*p*-value-based LD clumping is applied to select independent IVs for the BMI-BMI analysis, the confidence intervals (CIs) of MR-GMM, MR-Egger, and MR-APSS include the true causal effect *β* = 1 across almost all thresholds, although MR-Egger exhibits greater bias compared to MR-GMM and MR-APSS. This discrepancy arises because MR-GMM and MR-APSS explicitly account for the first-part selection bias caused by *p*-value thresholds using truncated normal models. In contrast, other methods tend to exclude the true causal effect from their CIs as the *p*-value threshold increases, as they fail to correct for the first-part selection bias in their models. Moreover, MR-GMM demonstrates shorter CI lengths compared to the other methods, suggesting its higher precision and reliability in these real-data scenarios.

**Fig 3:**
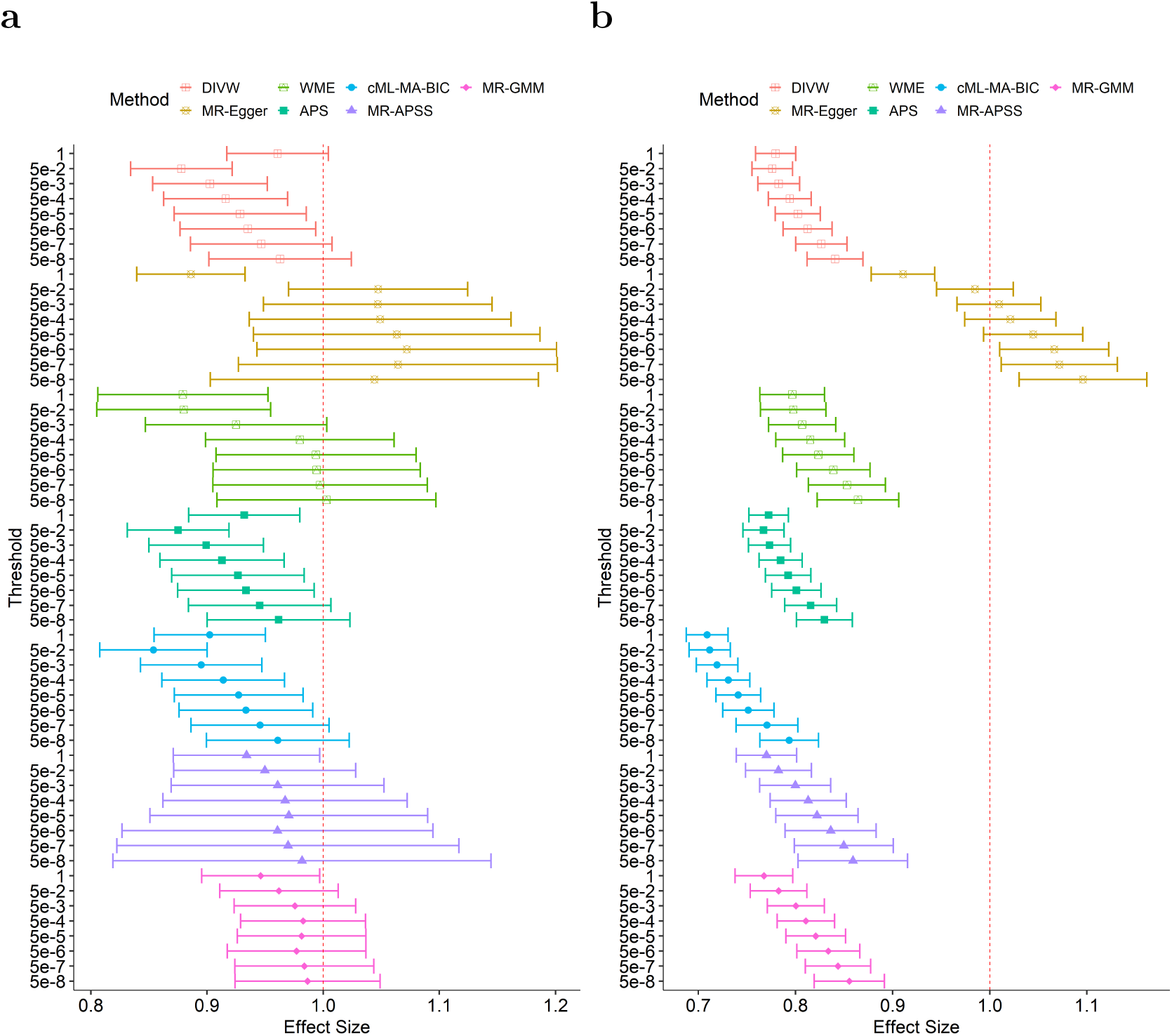
Benchmark BMI-BMI identity causal effect estimates obtained by using pseudo-*p*-value-based versus *p*-value-based LD clumping procedures under different IV selection thresholds. **a**. The forest plot of all methods using pseudo-*p*-value-based LD clumping procedure, which is conducted by first clumping based on pseudo *p*-values and then thresholding based on observed *p*-values. The 95% confidence intervals of point estimates are represented by error bars. **b**. The forest plot of all methods using standard *p*-value-based LD clumping procedure. These plots allow for a visual comparison of how different clumping methods and IV selection thresholds affect the estimation of causal effects in MR studies.

Figure 3b presents the results of the BMI-BMI analysis performed using the *p*-value-based LD clumping procedure. While the CI lengths are shorter under this approach, all methods, including MR-GMM and MR-APSS, exhibit severe biases, with the causal effect estimates systematically pulled toward 0. These biases intensify as the *p*-value threshold increases. This phenomenon arises due to the second-part selection bias introduced by the *p*-value-based LD clumping procedure, which preferentially selects the index SNPs with the smallest GWAS *p*-value. The results of the HDL-HDL analysis, illustrated in Figure S2, are consistent with those observed in the BMI-BMI analysis. By comparing the left and right panels of Figure 3 and Figure S2, it becomes evident that the pseudo-*p*-value-based LD clumping procedure substantially reduces selection bias in MR analyses, highlighting its advantage over *p*-value-based LD clumping.

### 2.4 Negative control study: The causal effects of complex traits on blonde hair color

We conducted a negative control study to assess the type I errors of the competing methods in real data scenarios. In this study, 81 complex traits were used as exposures, and blonde hair color before graying was chosen as the outcome as it was largely determined at birth and was not expected to be influenced by the selected exposures [19]. We further varied the criterion for selecting relevant IVs to validate the consistency of the results. Since not all complex traits exhibit high heritability, some traits may lack enough strongly associated SNPs after applying the pseudo-*p*-value-based LD clumping procedure. Therefore, we considered the following four selection criteria, aiming to include as many IVs as possible while maintaining a relatively high average IV strength: (i) SNPs with *p*-values less than 0.05; (ii) SNPs with *p*-values less than 0.1; (iii) the top 50 significant SNPs; and (iv) the top 500 significant SNPs. Criteria i and ii are the common thresholds in MR studies, and criteria iii and iv correspond to the simulation settings.

The quantile-quantile (Q-Q) plots of negative log-transformed *p*-values for the competing methods under all four selecting criteria are shown in Figure 4, with the genomic inflation factor, *λ*_GC_, for each method calculated and displayed in the top left corner of the plots [20]. Regardless of whether SNPs are selected based on *p*-value thresholds or rankings, cML-MA-BIC consistently fails to control type I error rates in the real data analyses. However, its performance improves when using selection criterion iii, as shown in Figure 4c. This observation aligns with findings from simulation studies, suggesting that the biases may be attributed to weak pleiotropic effects. Enhancing the average IV strength or perturbing the GWAS summary statistics could potentially improve cML-MA-BIC’s performance. Meanwhile, other methods demonstrate better control of type I error rates, although MR-Egger and MR-APSS show strong conservatism when applying selection criterion iv, as demonstrated in Figure 4d. This conservatism may be caused by the presence of a large number of weak instruments in the dataset. In a side by side comparison of MR-GMM under the four selection criteria, it exhibits the best control of type I error rates when IVs are selected using the *p*-value threshold of 0.05 (Figure 4a), as suggested by its genomic inflation factor, *λ*_MR-GMM_.

**Fig 4:**
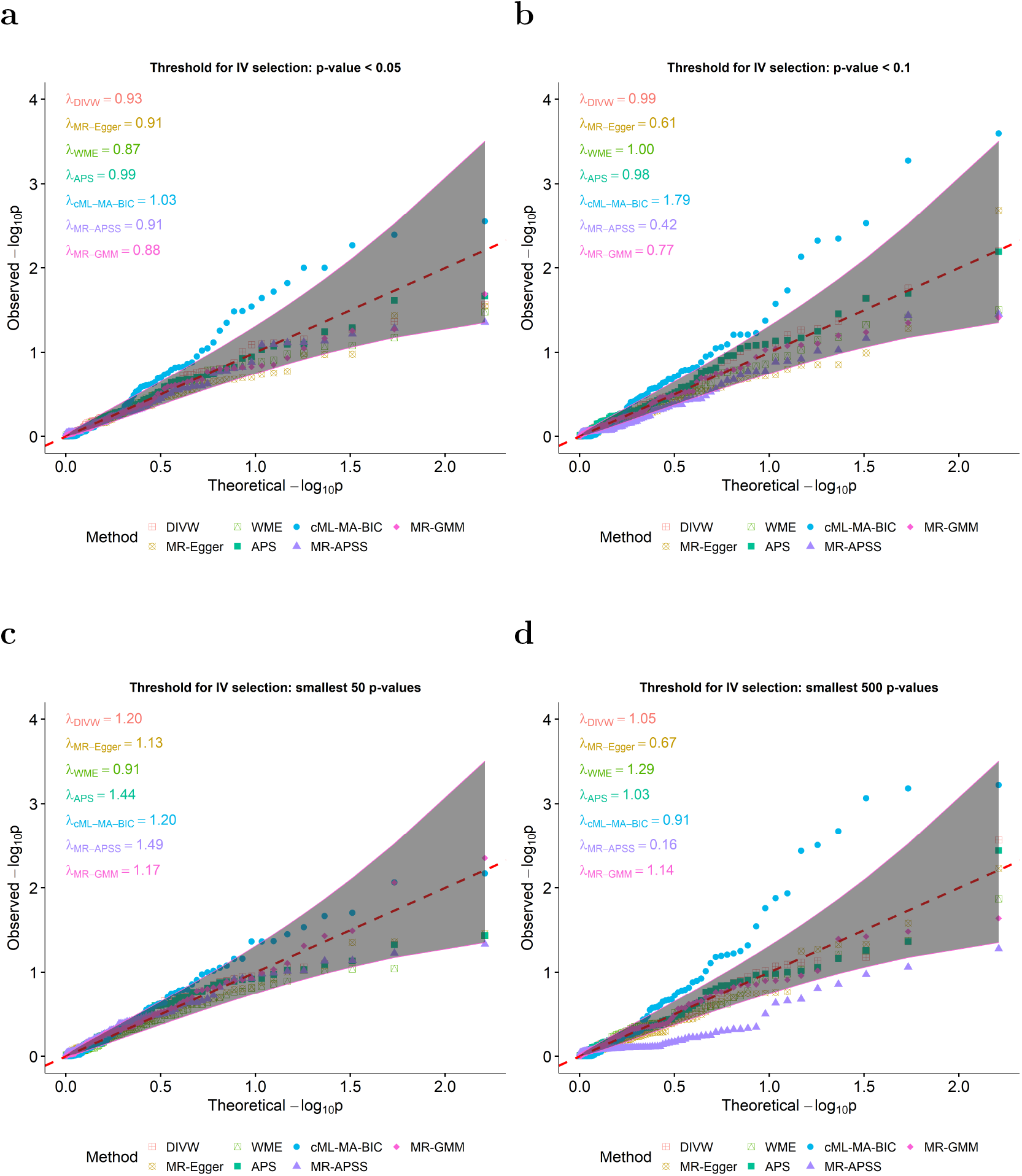
The ability of competing methods to control type I error rates in real data scenarios, illustrated through a negative control study. This study uses 81 different traits as exposures and blond hair color as the outcome. Q-Q plots of the negative log-transformed *p*-values for each method are presented, with the *λ*_GC_ values listed in the top-left corner of each plot for quantitative comparison. It is important to note that the variance of *λ*_GC_ can be large as *λ*_GC_ is calculated using 81 *χ*^2^ statistics. The grey regions on the plots represent the 95% confidence intervals for these *p*-values. After the pseudo-*p*-value-based LD clumping procedure, SNPs are selected based on different observed *p*-values: **a**. threshold of *p*-value *<* 0.05; **b**. threshold of *p*-value *<* 0.1; **c**. smallest 50 *p*-values **d**. smallest 500 *p*-values.

### 2.5 An application in proteome-wide MR identifies 45 CHD-related plasma proteins

Proteomics offers profound insights into the molecular mechanisms underlying diseases [21]. The plasma proteome encompasses a diverse array of proteins actively participating in various biological processes, which has made it a focal point of research [22]. Numerous studies have shown that plasma proteins are associated with heart diseases, and these proteins have the potential to serve as biomarkers for disease diagnosis, thereby facilitating drug development [21, 23, 24].

To further broaden our knowledge of CHD-related plasma proteins and provide clues for drug target, we applied MR-GMM to a proteome-wide MR analysis along with the other methods. Similar to Figure 4, the Q-Q plot of negative log-transformed *p*-values and the genomic inflation factors for each method are shown in Figure 5a. The IVs were selected using a *p*-value threshold of 0.05, as MR-GMM performs optimally under this threshold, as shown in Figure 4. The results under other selection criteria are presented in Figure S3. Consistent with the simulation results, MR-GMM found more significant proteins compared to other methods, while retaining a reasonable genomic inflation factor of *λ*_MR-GMM_ = 1.07. There are two main reasons for MR-GMM’s superior performance in identifying causal relationships. First, it enhances statistical power even when many weak or null IVs are involved in the analysis, as demonstrated in Figure 2. Second, it effectively accounts for the selection bias introduced by *p*-value thresholds used to select strong IVs, which, as noted earlier, tend to attenuate causal effects towards zero.

**Fig 5:**
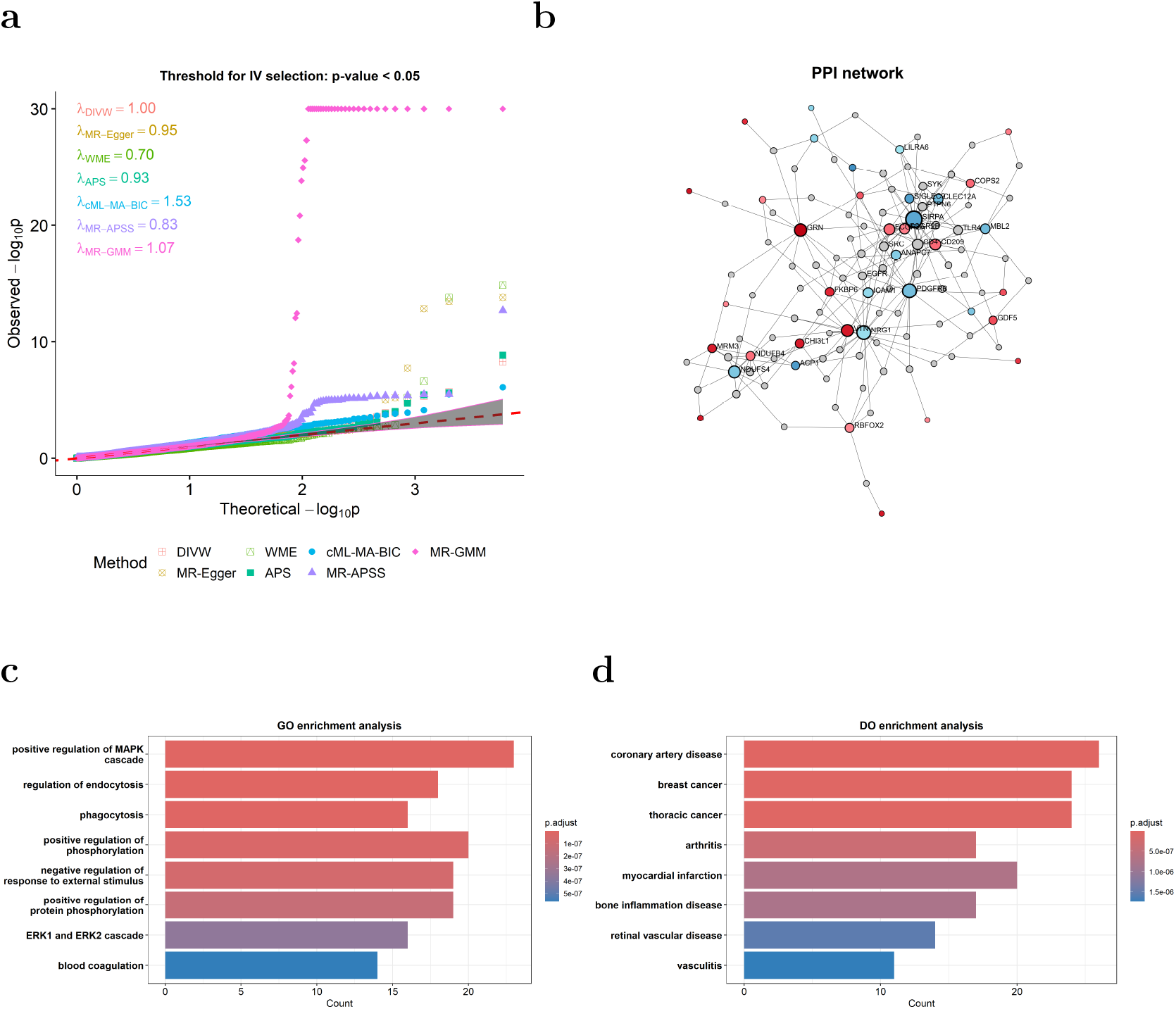
The application of MR-GMM in a proteome-wide MR study to identify plasma protein risk factors for CHD. **a**. Q-Q plots of the negative log-transformed *p*-values for each method, with the *λ*_GC_ values listed in the top-left corner of each plot. The grey regions represent the 95% confidence intervals for these *p*-values. The trimmed PPI network of 45 proteins identified by MR-GMM, with protein sizes proportional to their degrees. Proteins are color-coded according to their causal effects on CHD: red represents proteins that increase CHD risk, and blue indicates those that decrease it. **c**. The gene ontology enrichment analysis for biological processes using the proteins from the PPI network. The processes are ranked by significance, with the most significant ones listed at the top. **d**. The plot of disease ontology enrichment analysis using the proteins from the PPI network. The diseases are ranked by significance, with the most significant ones listed at the top.

Importantly, MR-GMM identifies 181 proteins with significant causal effects on CHD at the nominal level *α* = 0.05 (see Supplementary Data 2). To control the family-wise error rate, we employ the Holm-Bonferroni method at the nominal level *α* = 0.05 across 2992 tests, which is more powerful than the standard Bonferroni correction and does not rely on the independence of hypotheses [25]. Among the 45 proteins identified after correction (for details see Table S6), histo-blood group ABO system transferase (ABO) is a known protein causally relevant to CHD [26, 27]. Our results provide strong evidence that plasma ABO is associated with an increased risk of CHD (95% CI: [0.0326, 0.0329]). The ABO protein is the basis of the ABO blood group system. It can affect multiple aspects of von Willebrand factor (VWF) [28], a well-characterized biomarker of cardiovascular risk which promotes aggregation of platelets at high shear sites [29]. Genome-wide association studies have demonstrated that non-O ABO glycotransferase activity is related with coronary thrombosis, potentially mediated by ABO blood group carbohydrate modification of VWF [30].

We further investigated the network relationships among these proteins using NetworkAnalyst [31] with STRING database [32]. The trimmed protein-protein interaction (PPI) network is visualized in Figure 5b, with proteins color-coded to reflect their causal effects on CHD: red represents proteins that increase CHD risk, while blue indicates those that decrease it. Proteins were ranked by their network degrees (number of connections) and absolute values of their log-transformed z-scores (see Supplementary Data 3). The top three proteins in the network are tyrosine-protein phosphatase non-receptor type substrate 1 (SIRPA), platelet-derived growth factor receptor beta (PDGFRB), and neuregulin-1 (NRG1). Literatures have shown the protective roles of SIRPA, PDGFRB, NRG1 in CHD via pathways such as Toll-like receptor 4/nuclear factor-*κ*B, PDGFRB/EZH2, and NRG-1/ErbB [33–35], among others. Figures 5c and 5d represent subsequent gene ontology (GO) enrichment analysis [36, 37] of biological processes and disease ontology (DO) enrichment analysis [38] using 116 proteins from the PPI network with *clusterProfiler* [39] and *enrich-plot* [40]. Figure 5c reveals these proteins are involved in essential processes, including the regulation of MAPK cascade, endocytosis, and phosphorylation, which are known to be associated with cardiovascular diseases [41–44]. Additionally, Figure 5d high-lights enrichment in diseases such as coronary artery disease, breast cancer, and thoraric cancer. These analyses underscore the potential of these proteins as key players in both cardiovascular and broader systemic disease mechanisms. Finally, we conducted a network analysis to connect the 45 identified proteins with drugs in the DrugBank database [45]. The resulting network, shown in Figure S4, highlights interactions between these proteins and known drugs. Notably, abciximab, a monoclonal antibody used in the treatment of coronary artery disease [46], is associated with three proteins: low-affinity immunoglobulin gamma Fc region receptors II-a/II-b (FCGR2A/FCGR2B) and vitronectin (VTN). These proteins have been identified as CHD-related genes in prior studies [47, 48]. This finding provides valuable insights for advancing the development of targeted therapies for CHD.

## 3 Discussion

In this study, we proposed a novel MR method, MR-GMM, which leverages a GMM framework to estimate causal effects in the presence of weak IVs and pleiotropy. Unlike existing MR methods, MR-GMM explicitly classifies IVs into four categories—Invalid, Valid, Invalid&Null, and Null IVs—allowing for greater flexibility and robustness in real-world applications where assumptions such as no pleiotropy or strong instruments may not hold. By incorporating a broad set of IVs, MR-GMM enhances statistical power in scenarios where exposures have relatively low heritability. Additionally, MR-GMM accounts for selection bias caused by *p*-value thresholds used to select SNPs associated with exposures. Since this bias typically attenuates causal effect estimates toward zero, the correction introduced by MR-GMM will further increase the power to detect causal effects. MR-GMM outperforms existing methods in terms of type I error, statistical power, bias, and precision, as demonstrated through simulation studies.

Importantly, MR-GMM proposes a refined pseudo-*p*-value-based LD clumping pro-cedure to mitigate selection bias inherent in the traditional *p*-value-based LD clumping. This refined approach was validated through analyses estimating identity causal effects and a negative control study, demonstrating substantial improvements in the reliability of causal effect estimates.

Nevertheless, MR-GMM has several limitations that require further refinement. First, as shown in Figures S5 and S6, MR-GMM struggles to control type I error rates when the number of IVs is too small or when the average IV strength is too weak, although under these cases the point estimate is still unbiased. This issue may arise because the variance estimation in MR-GMM relies on large-sample properties, and with a limited number of IVs, the asymptotic of MR-GMM can deviate from its actual distribution. Additionally, the asymptotic properties of MR-GMM depend on certain regularity conditions, which may be violated when the average IV strength is insufficient. This issue suggests that either a larger number of SNPs should be used as IVs, or SNPs with higher average IV strength should be prioritized. The biased estimation of SE may potentially be addressed by resampling strategies, such as bootstrapping or perturbation, which can improve the variance estimation in small-sample scenarios. Second, MR-GMM only considers independent SNPs in the model. This could result in a loss of statistical power, for including more SNPs in LD with strong IVs could potentially improve both the power and precision in MR analyses [49, 50]. Lastly, although we proposed the pseudo-*p*-value-based LD clumping procedure to mitigate selection bias, a standardized method for determining the *p*-value threshold for selecting SNPs associated with exposures remains necessary. In this work, we explored four different selection criteria and identified the optimal one through negative control studies, but establishing a universal standard would enhance the robustness and generalizability of the approach.

From a practical perspective, the successful application of MR-GMM in proteome-wide MR analysis underscores its utility in uncovering novel causal relationships. The identification of 45 CHD-related proteins, including well-known protective proteins such as SIRPA, PDGFRB, and NRG1, illustrates its capacity to provide biologically meaningful insights. Furthermore, combining the results of MR-GMM with protein-drug interaction networks, as demonstrated by the identification of abciximab’s associations with three CHD-related proteins, highlights its translational potential in drug discovery and precision medicine.

## 4 Methods

### 4.1 MR-GMM model for four classes of IVs

MR-GMM takes the effect size estimates and their SEs from the exposure and outcome GWAS summary statistics as input. Specifically, let 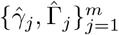 denote the estimated effects of the SNP *G*_*j*_ on the exposure *X* and outcome *Y*, respectively, with corresponding SEs, 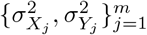, obtained from marginal regression of *X* and *Y* on *G*_*j*_. Following the measurement error model proposed by RAPS [5], we assume that the observed effect sizes 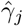 and 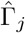 follow normal distributions around their true effect sizes 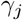 and 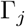 with variances 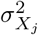 and 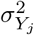, respectively, formally,

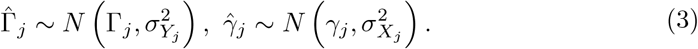

We model the true effect sizes *γ*_*j*_ and Γ_*j*_ with a two dimensional spike-and-slab prior as described in Equation (1). To identify the causal effect from the model, we assume that the latent effects *γ*_*j*0_ and *α*_*j*0_ follow independent normal distributions, *N* (0, *η*^*−*1^), *N* (0, *τ* ^*−*1^), respectively. Further, the binary indicators (*Z*_*γ*_, *Z*_*α*_) are re-encoded into independent and identically distributed (i.i.d.) multinomial random variables, *L*_*j*_, for *j* = 1, …, *m*, as follows,

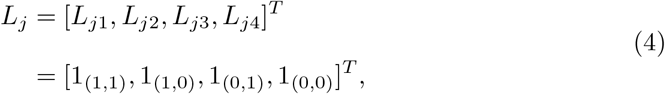

Where 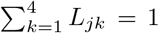, and 1_(*a,b*)_ is an indicator function equal to 1 if (*Z*_*γ j*_, *Z*_*α j*_) = (*a, b*), and 0 otherwise. Then the index *k* corresponds to *L*_*jk*_ = 1 aligns with the *k*th class of SNPs. Let *π*_*k*_ = *P* (*L*_*jk*_ = 1) denotes the prior probability of the *k*th class, and *π* = [*π*_1_, *π*_2_, *π*_3_, *π*_4_]^*T*^ is the vector of prior probabilities. The marginal likelihood function of the model after integrating out *γ*_*j*0_, *α*_*j*0_, *L*_*j*_ is given by

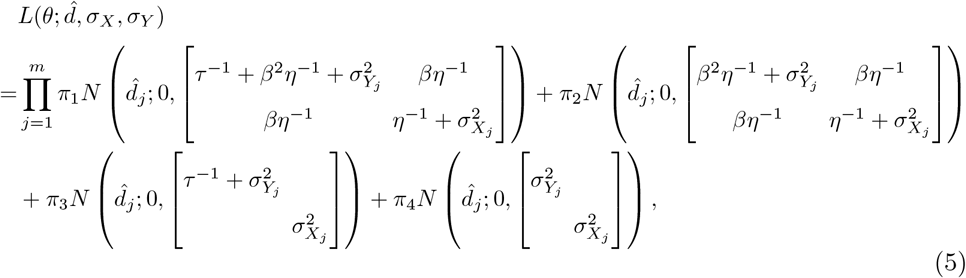

where *θ* = (*β, τ, η, π*^*T*^)^*T*^ are parameters to be estimated, and 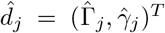 are observed data for effect sizes. We further penalize the likelihood function in Equation (5) to constrain the probabilities *π*_*k*_ by incorporating the following penalty term:

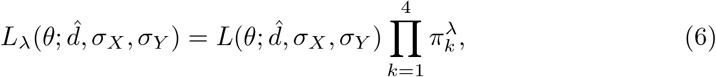

where *λ* are hyperparameters representing prior knowledge about the least number of SNPs in each class. In this study, we set 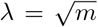, which controlled type I error rates well in real data scenarios. To optimize the penalized likelihood function efficiently and robustly, we utilize the deterministic annealing expectation-maximization (DAEM) algorithm, a variation of the traditional EM algorithm [51]. The DAEM algorithm mitigates issues related to convergence to local optima by gradually introducing randomness through annealing [52].

Under certain regularity conditions, 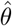, the MLE of *θ* is consistent and asymptotically normal, i.e.,

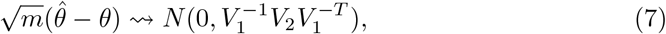

where 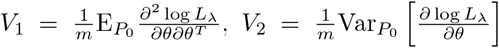, and *P*_0_ stands for the distribution of observed data under true parameters. The variance-covariance matrix can be estimated by a Sandwich estimator. The details of the DAEM algorithm and variance-covariance matrix are provided in the Supplementary Note.

### 4.2 Pseudo-*p*-value-based LD clumping procedure

To mitigate the selection bias introduced by retaining the SNP with the smallest GWAS-reported *p*-value as the index SNP within a chromosome region in the *p*-value-based LD clumping procedure, we propose using pseudo-*p*-values, calculated using the equation below, as substitutes for the observed *p*-values,

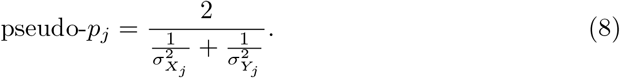

The pseudo-*p*-value represents the pooled measurement error of the *j*-th SNP. Actually, the variance estimate of the effect size for the *j*-th SNP in GWAS summary data is closely related to the sample size and can be approximately treated as the inverse of the sample size [53]. Consequently, the pseudo-*p*-values derived from this equation can also be interpreted as the inverse of the average sample size of the exposure and outcome datasets.

Unlike the traditional *p*-value-based LD clumping procedure, the pseudo-*p*-value-based LD clumping procedure selects the SNP with the smallest pseudo-*p*-value as the index SNP (still implemented with *plink* [54]). This approach achieves two key advantages: (i) it prioritizes SNPs with the largest pooled sample sizes, thereby reducing the impact of measurement errors, and (ii) it mitigates selection bias caused by *p*-value ranking. By addressing these issues, the pseudo-*p*-value-based procedure improves the reliability and accuracy of SNP selection in MR analyses.

The detailed information of the BMI and HDL datasets used to compare the two LD clumping procudures is provided in Tables S1–S4.

### 4.3 Data generating process for simulation studies

We generated the summary statistics based on the measurement error model (3). To be specific, we first generate the indicator *L*_*j*_ for SNP *j* from a multinomial distribution with probabilities *π*. Next, we generate the i.i.d. latent effects *γ*_*j*0_ and *α*_*j*0_ from normal distributions *N* (0, 10^*−*4^). The true effect sizes *γ*_*j*_ and Γ_*j*_ were then computed based on the latent effects and the indicators as follows,

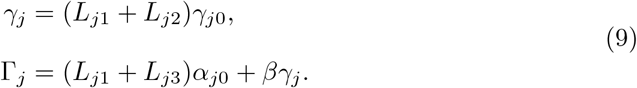

The observed effect sizes 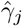 and 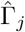 were generated by adding Gaussian noise to the true effect sizes, with variances 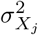 and 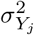 obtained from the *bmi*.*bmi* dataset included in the *mr*.*raps* package [5], respectively.

The four scenarios were generated as follows:

- Balanced: *π* = [0.25, 0.25, 0.25, 0.25]^*T*^ ;
- Valid: *π* = [0.1, 0.1, 0.1, 0.7]^*T*^ ;
- Invalid & Null: *π* = [0.1, 0.1, 0.7, 0.1]^*T*^ ;
- Null: *π* = [0.1, 0.1, 0.1, 0.7]^*T*^.

Without loss of generality, the variances 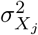 and 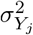 were taken from the first 500 rows of the *bmi*.*bmi* dataset. The causal effect size was varied from −0.15 to 0.15 in increments of 0.05 across all scenarios, with powers evaluated using 1000 simulated replicates.

### 4.4 GWAS data preprocessing

#### 4.4.1 Choice of exposures

For the negative control study, to select as many exposures as possible, we used all exposures from the MRC IEU OpenGWAS data infrastructure whose OpenGWAS IDs beginning with “ieu-a-” [55]. To ensure compatibility with the outcome and avoid population structure biases, we selected exposures based on individuals of European ancestry and those that included both male and female participants. Since the outcome were sourced from the UK BioBank Consortium [56], we further excluded exposures sourced from UK BioBank or MRC-IEU Consortium and filtered for exposures with sample sizes greater than 10,000 to reduce the influence of sample overlap and measurement errors. Additionally, to prevent redundancies, we removed exposures categorized under the subcategory *cancer* and deduplicated the remaining exposures.

For the proteome-wide MR analysis, we used plasma proteins from a proteome-wide genomic atlas as exoposures (OpenGWAS IDs beginning with “prot-a-”) [57]. After removing duplicated proteins from the original 3284 exposures, a total of 2992 unique proteins were retained for the analysis.

For convenience, we collected all GWAS datasets from the MRC IEU OpenG-WAS data infrastructure, which were processed in variant call format (VCF) [58]. The detailed information of all exposures and outcomes used in the negative control study and proteome-wide MR analysis is provided in Table S5 (see Supplementary Data 1 for full results) and Supplementary Data 2, respectively.

#### 4.4.2 Quality control for GWAS summary statistics

To further reduce biases in MR analysis, we implemented the following quality control procedures for the GWAS summary statistics: (i) extracted SNPs common to both the exposure and outcome datasets; (ii) join biallelic sites into multiallelic records and removed multiallelic variants, palindromic SNPs, and SNPs with a minor allele frequency (MAF) less than 0.01; (iii) applied the pseudo-*p*-value-based LD clumping procedure to select independent variants with *r*^2^ *<* 0.001 and a window size of 10,000 Kb, as suggested by the *TwoSampleMR* package [59]; (iv) harmonized the exposure and outcome datasets using the *harmonize* function from the *TwoSam-pleMR* package, which produced a merged dataset. The *p*-value threshold for selecting strong IVs was applied to this merged dataset. All quality control procedures were implemented using *bcftools* [60].

### 4.5 Accounting for selection bias caused by the selection of strong IVs

We propose using a truncated GMM to account for selection bias caused by the selection of strong IVs. Specifically, let S_*t*_≠∅ denote selection rule

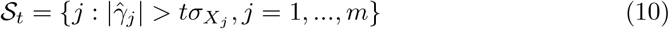

corresponding to selecting SNPs using a *p*-value threshold of 2Φ(−*t*), with Φ representing the cumulative distribution function of the standard normal distribution. Then the corresponding truncated GMM can be derived as,

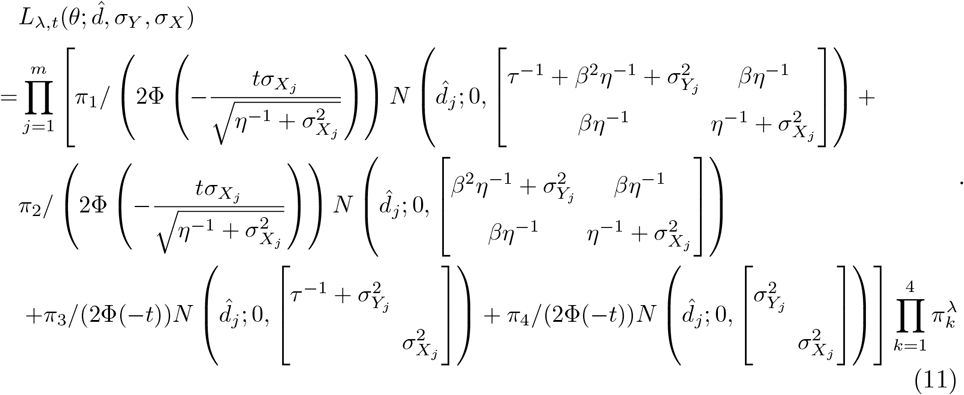

The optimization of Equation (11) was performed also using the DAEM algorithm described in the Supplementary Note.

## 5 Data availability

The reference LD panel is available to be downloaded from https://github.com/perslab/CELLECT/tree/master/data/ldsc/1000G EUR Phase3 plink. All GWAS summary statistics used can be accessed from the MRC IEU OpenGWAS data infrastructure (https://gwas.mrcieu.ac.uk/datasets/).

## 6 Code availability

The implementation of MR-GMM is publicly available at https://github.com/YuquanW/mr.gmm. Additionally, the code for the pseudo-*p*-value-based LD clumping procedure, along with resources for reproducibility, can be found at https://github.com/YuquanW/Clumping.

## 8 Acknowledgements

This research was supported by National Key R&D Program of China [2023YFF1205101].

## 9 Author contributions

Y-Q.H. provided funding support. Y.W. conceptualized the study design and developed the statistical methodology with assistance from Y.C., D.C., and D.S. Y.W. conducted the data analyses wrote the first draft of manuscript with assistance from L.F. S.S., A.C., and Y-Q.H. provided reviews to refine the manuscript and approved the final version.

## 10 Competing interests

The authors declare that the research was conducted in the absence of any commercial or financial relationships that could be construed as a potential conflict of interest.

## Notes

### Competing Interest Statement

The authors have declared no competing interest.

### Author Declarations

All source data used in this study were openly accessible prior to the initiation of the research. These data can be obtained from the following sources: 1. MRC IEU OpenGWAS data infrastructure: Genome-wide association summary statistics for 2992 plasma proteins, 81 common traits, blonde hair color, and coronary heart disease. The datasets are available at https://gwas.mrcieu.ac.uk/datasets/. 2.Linkage disequilibrium reference panel. The dataset is available at https://github.com/perslab/CELLECT/blob/master/data/ldsc/1000G_EUR_Phase3_plink/.

