## Supplementary Materials for "Pseudo-*p*-Value-Based Clumping Enhanced Proteome-wide Mendelian Randomization with Application in Identifying Coronary Heart Disease-Associated Plasma Proteins"

#### 1 Supplementary Notes

We organize the supplementary notes as follows: In subsection **??**, we provide the variance-covariance matrix for the MR-GMM estimator. In subsection **??**, we outline the steps of the Deterministic Annealing Expectation-Maximization (DAEM) algorithm, including its optimization process presented in Algorithm 1.

#### 1.1 The variance-covariance matrix of the MR-GMM estimator

The derivation of the variance-covariance matrix of the MR-GMM estimator is omitted for brevity. We only provide the final expression here. One can verify the results using the R package *NumDeriv*. In practice, because we only need the variance of $\hat{\beta}$ for subsequential analyses, we reparameterize the original parameter $\theta$ as $\tilde{\theta} = [\beta, \tilde{\tau}, \tilde{\eta}, \tilde{\pi}_1, \tilde{\pi}_2, \tilde{\pi}_3]^T = \left[\beta, \log\tau, \log\eta, \log\frac{\pi_1}{1-\sum_{k=1}^{3}\pi_k}, \log\frac{\pi_2}{1-\sum_{k=1}^{3}\pi_k}, \log\frac{\pi_3}{1-\sum_{k=1}^{3}\pi_k}\right]^T$ to avoid calculation problems arising from large values of $\tau$ and $\eta$. Let

$$
\begin{aligned}
\tilde{V}_1 =& \frac{1}{m}\mathrm{E}_{P_0}\frac{\partial^2 \log L_\lambda}{\partial\tilde{\theta}\partial\tilde{\theta}^T} \\
=& \frac{1}{m}\sum_{j=1}^{m}\mathrm{E}_{P_0}\begin{bmatrix}
\frac{\partial^2 l_j^\lambda}{\partial\beta^2} & \frac{\partial^2 l_j^\lambda}{\partial\beta\partial\tilde{\tau}} & \frac{\partial^2 l_j^\lambda}{\partial\beta\partial\tilde{\eta}} & \frac{\partial^2 l_j^\lambda}{\partial\beta\partial\tilde{\pi}_1} & \frac{\partial^2 l_j^\lambda}{\partial\beta\partial\tilde{\pi}_2} & \frac{\partial^2 l_j^\lambda}{\partial\beta\partial\tilde{\pi}_3} \\
\frac{\partial^2 l_j^\lambda}{\partial\tilde{\tau}\partial\beta} & \frac{\partial^2 l_j^\lambda}{\partial\tilde{\tau}^2} & \frac{\partial^2 l_j^\lambda}{\partial\tilde{\tau}\partial\tilde{\eta}} & \frac{\partial^2 l_j^\lambda}{\partial\tilde{\tau}\partial\tilde{\pi}_1} & \frac{\partial^2 l_j^\lambda}{\partial\tilde{\tau}\partial\tilde{\pi}_2} & \frac{\partial^2 l_j^\lambda}{\partial\tilde{\tau}\partial\tilde{\pi}_3} \\
\frac{\partial^2 l_j^\lambda}{\partial\tilde{\eta}\partial\beta} & \frac{\partial^2 l_j^\lambda}{\partial\tilde{\eta}\partial\tilde{\tau}} & \frac{\partial^2 l_j^\lambda}{\partial\tilde{\eta}^2} & \frac{\partial^2 l_j^\lambda}{\partial\tilde{\eta}\partial\tilde{\pi}_1} & \frac{\partial^2 l_j^\lambda}{\partial\tilde{\eta}\partial\tilde{\pi}_2} & \frac{\partial^2 l_j^\lambda}{\partial\tilde{\eta}\partial\tilde{\pi}_3} \\
\frac{\partial^2 l_j^\lambda}{\partial\tilde{\pi}_1\partial\beta} & \frac{\partial^2 l_j^\lambda}{\partial\tilde{\pi}_1\partial\tilde{\tau}} & \frac{\partial^2 l_j^\lambda}{\partial\tilde{\pi}_1\partial\tilde{\eta}} & \frac{\partial^2 l_j^\lambda}{\partial\tilde{\pi}_1^2} & \frac{\partial^2 l_j^\lambda}{\partial\tilde{\pi}_1\partial\tilde{\pi}_2} & \frac{\partial^2 l_j^\lambda}{\partial\tilde{\pi}_1\partial\tilde{\pi}_3} \\
\frac{\partial^2 l_j^\lambda}{\partial\tilde{\pi}_2\partial\beta} & \frac{\partial^2 l_j^\lambda}{\partial\tilde{\pi}_2\partial\tilde{\tau}} & \frac{\partial^2 l_j^\lambda}{\partial\tilde{\pi}_2\partial\tilde{\eta}} & \frac{\partial^2 l_j^\lambda}{\partial\tilde{\pi}_2\partial\tilde{\pi}_1} & \frac{\partial^2 l_j^\lambda}{\partial\tilde{\pi}_2^2} & \frac{\partial^2 l_j^\lambda}{\partial\tilde{\pi}_2\partial\tilde{\pi}_3} \\
\frac{\partial^2 l_j^\lambda}{\partial\tilde{\pi}_3\partial\beta} & \frac{\partial^2 l_j^\lambda}{\partial\tilde{\pi}_3\partial\tilde{\tau}} & \frac{\partial^2 l_j^\lambda}{\partial\tilde{\pi}_3\partial\tilde{\eta}} & \frac{\partial^2 l_j^\lambda}{\partial\tilde{\pi}_3\partial\tilde{\pi}_1} & \frac{\partial^2 l_j^\lambda}{\partial\tilde{\pi}_3\partial\tilde{\pi}_2} & \frac{\partial^2 l_j^\lambda}{\partial\tilde{\pi}_3^2}
\end{bmatrix}, \\
\tilde{V}_2 =& \frac{1}{m}\mathrm{Var}_{P_0}\left[\frac{\partial \log L_\lambda}{\partial\tilde{\theta}}\right] \\
=& \frac{1}{m}\sum_{j=1}^{m}\mathrm{E}_{P_0}\begin{bmatrix}
(\frac{\partial l_j^\lambda}{\partial\beta})^2 & \frac{\partial l_j^\lambda}{\partial\beta}\frac{\partial l_j^\lambda}{\partial\tilde{\tau}} & \frac{\partial l_j^\lambda}{\partial\beta}\frac{\partial l_j^\lambda}{\partial\tilde{\eta}} & \frac{\partial l_j^\lambda}{\partial\beta}\frac{\partial l_j^\lambda}{\partial\tilde{\pi}_1} & \frac{\partial l_j^\lambda}{\partial\beta}\frac{\partial l_j^\lambda}{\partial\tilde{\pi}_2} & \frac{\partial l_j^\lambda}{\partial\beta}\frac{\partial l_j^\lambda}{\partial\tilde{\pi}_3} \\
\frac{\partial l_j^\lambda}{\partial\tilde{\tau}}\frac{\partial l_j^\lambda}{\partial\beta} & (\frac{\partial l_j^\lambda}{\partial\tilde{\tau}})^2 & \frac{\partial l_j^\lambda}{\partial\tilde{\tau}}\frac{\partial l_j^\lambda}{\partial\tilde{\eta}} & \frac{\partial l_j^\lambda}{\partial\tilde{\tau}}\frac{\partial l_j^\lambda}{\partial\tilde{\pi}_1} & \frac{\partial l_j^\lambda}{\partial\tilde{\tau}}\frac{\partial l_j^\lambda}{\partial\tilde{\pi}_2} & \frac{\partial l_j^\lambda}{\partial\tilde{\tau}}\frac{\partial l_j^\lambda}{\partial\tilde{\pi}_3} \\
\frac{\partial l_j^\lambda}{\partial\tilde{\eta}}\frac{\partial l_j^\lambda}{\partial\beta} & \frac{\partial l_j^\lambda}{\partial\tilde{\eta}}\frac{\partial l_j^\lambda}{\partial\tilde{\tau}} & (\frac{\partial l_j^\lambda}{\partial\tilde{\eta}})^2 & \frac{\partial l_j^\lambda}{\partial\tilde{\eta}}\frac{\partial l_j^\lambda}{\partial\tilde{\pi}_1} & \frac{\partial l_j^\lambda}{\partial\tilde{\eta}}\frac{\partial l_j^\lambda}{\partial\tilde{\pi}_2} & \frac{\partial l_j^\lambda}{\partial\tilde{\eta}}\frac{\partial l_j^\lambda}{\partial\tilde{\pi}_3} \\
\frac{\partial l_j^\lambda}{\partial\tilde{\pi}_1}\frac{\partial l_j^\lambda}{\partial\beta} & \frac{\partial l_j^\lambda}{\partial\tilde{\pi}_1}\frac{\partial l_j^\lambda}{\partial\tilde{\tau}} & \frac{\partial l_j^\lambda}{\partial\tilde{\pi}_1}\frac{\partial l_j^\lambda}{\partial\tilde{\eta}} & (\frac{\partial l_j^\lambda}{\partial\tilde{\pi}_1})^2 & \frac{\partial l_j^\lambda}{\partial\tilde{\pi}_1}\frac{\partial l_j^\lambda}{\partial\tilde{\pi}_2} & \frac{\partial l_j^\lambda}{\partial\tilde{\pi}_1}\frac{\partial l_j^\lambda}{\partial\tilde{\pi}_3} \\
\frac{\partial l_j^\lambda}{\partial\tilde{\pi}_2}\frac{\partial l_j^\lambda}{\partial\beta} & \frac{\partial l_j^\lambda}{\partial\tilde{\pi}_2}\frac{\partial l_j^\lambda}{\partial\tilde{\tau}} & \frac{\partial l_j^\lambda}{\partial\tilde{\pi}_2}\frac{\partial l_j^\lambda}{\partial\tilde{\eta}} & \frac{\partial l_j^\lambda}{\partial\tilde{\pi}_2}\frac{\partial l_j^\lambda}{\partial\tilde{\pi}_1} & (\frac{\partial l_j^\lambda}{\partial\tilde{\pi}_2})^2 & \frac{\partial l_j^\lambda}{\partial\tilde{\pi}_2}\frac{\partial l_j^\lambda}{\partial\tilde{\pi}_3} \\
\frac{\partial l_j^\lambda}{\partial\tilde{\pi}_3}\frac{\partial l_j^\lambda}{\partial\beta} & \frac{\partial l_j^\lambda}{\partial\tilde{\pi}_3}\frac{\partial l_j^\lambda}{\partial\tilde{\tau}} & \frac{\partial l_j^\lambda}{\partial\tilde{\pi}_3}\frac{\partial l_j^\lambda}{\partial\tilde{\eta}} & \frac{\partial l_j^\lambda}{\partial\tilde{\pi}_3}\frac{\partial l_j^\lambda}{\partial\tilde{\pi}_1} & \frac{\partial l_j^\lambda}{\partial\tilde{\pi}_3}\frac{\partial l_j^\lambda}{\partial\tilde{\pi}_2} & (\frac{\partial l_j^\lambda}{\partial\tilde{\pi}_3})^2
\end{bmatrix}.
\end{aligned} \tag{S1}
$$

Therefore, to obtain the variance of $\hat{\beta}$, it suffices to derive the first and second partial derivations of the individual penalized log-likelihood function $l_j^\lambda$ with respect to each element in $\tilde{\theta}$. They can be calculated as follows:

$$
\frac{\partial l_j^\lambda}{\partial\beta} = \frac{1}{L_j}\left(\frac{\pi_1 f_1}{\Phi\left(-\frac{t\sigma_{X_j}}{\sqrt{\eta^{-1}+\sigma_{X_j}^2}}\right)}\frac{\partial \log f_1}{\partial\beta} + \frac{\pi_2 f_2}{\Phi\left(-\frac{t\sigma_{X_j}}{\sqrt{\eta^{-1}+\sigma_{X_j}^2}}\right)}\frac{\partial \log f_2}{\partial\beta}\right)
$$

$$\frac{\partial l_j^\lambda}{\partial \tilde{\tau}} = \frac{\tau}{L_j}\left(\frac{\pi_1 f_1}{\Phi\left(-\frac{t\sigma_{X_j}}{\sqrt{\eta^{-1}+\sigma_{X_j}^2}}\right)}\frac{\partial \log f_1}{\partial \tau} + \frac{\pi_3 f_3}{\Phi\left(-t\right)}\frac{\partial \log f_3}{\partial \tau}\right)$$

$$\frac{\partial l_j^\lambda}{\partial \tilde{\eta}} = \frac{\eta}{L_j}\left(\frac{\pi_1 f_1}{\Phi\left(-\frac{t\sigma_{X_j}}{\sqrt{\eta^{-1}+\sigma_{X_j}^2}}\right)}\left(\frac{\partial \log f_1}{\partial \eta} - \frac{\partial \log \Phi\left(-\frac{t\sigma_{X_j}}{\sqrt{\eta^{-1}+\sigma_{X_j}^2}}\right)}{\partial \eta}\right) + \frac{\pi_2 f_2}{\Phi\left(-t\right)}\left(\frac{\partial \log f_2}{\partial \eta} - \frac{\partial \log \Phi\left(-\frac{t\sigma_{X_j}}{\sqrt{\eta^{-1}+\sigma_{X_j}^2}}\right)}{\partial \eta}\right)\right)$$

$$\frac{\partial l_j^\lambda}{\partial \tilde{\pi}_1} = \pi_1\left(\frac{f_1}{L_j\Phi\left(-\frac{t\sigma_{X_j}}{\sqrt{\eta^{-1}+\sigma_{X_j}^2}}\right)} - 1\right) + \frac{\lambda}{m}(1-4\pi_1)$$

$$\frac{\partial l_j^\lambda}{\partial \tilde{\pi}_2} = \pi_2\left(\frac{f_2}{L_j\Phi\left(-\frac{t\sigma_{X_j}}{\sqrt{\eta^{-1}+\sigma_{X_j}^2}}\right)} - 1\right) + \frac{\lambda}{m}(1-4\pi_2)$$

$$\frac{\partial l_j^\lambda}{\partial \tilde{\pi}_3} = \pi_3\left(\frac{f_3}{L_j\Phi\left(-t\right)} - 1\right) + \frac{\lambda}{m}(1-4\pi_3)$$

$$\frac{\partial^2 l_j^\lambda}{\partial \beta^2} = -\left(\frac{\partial l_j^\lambda}{\partial \beta}\right)^2 + \frac{1}{L_j}\left(\frac{\pi_1 f_1}{\Phi\left(-\frac{t\sigma_{X_j}}{\sqrt{\eta^{-1}+\sigma_{X_j}^2}}\right)}\left(\left(\frac{\partial \log f_1}{\partial \beta}\right)^2 + \frac{\partial^2 \log f_1}{\partial \beta^2}\right) + \frac{\pi_2 f_2}{\Phi\left(-\frac{t\sigma_{X_j}}{\sqrt{\eta^{-1}+\sigma_{X_j}^2}}\right)}\left(\left(\frac{\partial \log f_2}{\partial \beta}\right)^2 + \frac{\partial^2 \log f_2}{\partial \beta^2}\right)\right)$$

$$\frac{\partial^2 l_j^\lambda}{\partial \beta \partial \tilde{\tau}} = -\frac{\partial l_j^\lambda}{\partial \beta}\frac{\partial l_j^\lambda}{\partial \tilde{\tau}} + \frac{\tau \pi_1 f_1}{L_j\Phi\left(-\frac{t\sigma_{X_j}}{\sqrt{\eta^{-1}+\sigma_{X_j}^2}}\right)}\left(\frac{\partial \log f_1}{\partial \beta}\frac{\partial \log f_1}{\partial \tau} + \frac{\partial^2 \log f_1}{\partial \beta \partial \tau}\right)$$

$$\frac{\partial^2 l_j^\lambda}{\partial\beta\partial\tilde\eta} = -\frac{\partial l_j^\lambda}{\partial\beta}\frac{\partial l_j^\lambda}{\partial\tilde\eta} + \frac{\eta}{L_j}\left(\frac{\pi_1 f_1}{\Phi\left(-\frac{t\sigma_{X_j}}{\sqrt{\eta^{-1}+\sigma^2_{X_j}}}\right)}\left(\frac{\partial\log f_1}{\partial\beta}\left(\frac{\partial\log f_1}{\partial\eta} - \frac{\partial\log\Phi\left(-\frac{t\sigma_{X_j}}{\sqrt{\eta^{-1}+\sigma^2_{X_j}}}\right)}{\partial\eta}\right) + \frac{\partial^2\log f_1}{\partial\beta\partial\eta}\right)\right.$$

$$\left.\frac{\pi_2 f_2}{\Phi\left(-\frac{t\sigma_{X_j}}{\sqrt{\eta^{-1}+\sigma^2_{X_j}}}\right)}\left(\frac{\partial\log f_2}{\partial\beta}\left(\frac{\partial\log f_2}{\partial\eta} - \frac{\partial\log\Phi\left(-\frac{t\sigma_{X_j}}{\sqrt{\eta^{-1}+\sigma^2_{X_j}}}\right)}{\partial\eta}\right) + \frac{\partial^2\log f_2}{\partial\beta\partial\eta}\right)\right)$$

$$\frac{\partial^2 l_j^\lambda}{\partial\beta\partial\tilde\pi_1} = -\frac{\partial l_j^\lambda}{\partial\beta}\frac{\partial l_j^\lambda}{\partial\tilde\pi_1} + \frac{1}{L_j}\left(\frac{\pi_1(1-\pi_1)f_1}{\Phi\left(-\frac{t\sigma_{X_j}}{\sqrt{\eta^{-1}+\sigma^2_{X_j}}}\right)}\frac{\partial\log f_1}{\partial\beta} - \frac{\pi_1\pi_2 f_2}{\Phi\left(-\frac{t\sigma_{X_j}}{\sqrt{\eta^{-1}+\sigma^2_{X_j}}}\right)}\frac{\partial\log f_2}{\partial\beta}\right)$$

$$\frac{\partial^2 l_j^\lambda}{\partial\beta\partial\tilde\pi_2} = -\frac{\partial l_j^\lambda}{\partial\beta}\frac{\partial l_j^\lambda}{\partial\tilde\pi_2} + \frac{1}{L_j}\left(\frac{\pi_2(1-\pi_2)f_2}{\Phi\left(-\frac{t\sigma_{X_j}}{\sqrt{\eta^{-1}+\sigma^2_{X_j}}}\right)}\frac{\partial\log f_2}{\partial\beta} - \frac{\pi_1\pi_2 f_1}{\Phi\left(-\frac{t\sigma_{X_j}}{\sqrt{\eta^{-1}+\sigma^2_{X_j}}}\right)}\frac{\partial\log f_1}{\partial\beta}\right)$$

$$\frac{\partial^2 l_j^\lambda}{\partial\beta\partial\tilde\pi_3} = -\frac{\partial l_j^\lambda}{\partial\beta}\frac{\partial l_j^\lambda}{\partial\tilde\pi_3} + \frac{1}{L_j}\left(-\frac{\pi_1\pi_3 f_1}{\Phi\left(-\frac{t\sigma_{X_j}}{\sqrt{\eta^{-1}+\sigma^2_{X_j}}}\right)}\frac{\partial\log f_1}{\partial\beta} - \frac{\pi_2\pi_3 f_2}{\Phi\left(-\frac{t\sigma_{X_j}}{\sqrt{\eta^{-1}+\sigma^2_{X_j}}}\right)}\frac{\partial\log f_2}{\partial\beta}\right)$$

$$\frac{\partial^2 l_j^\lambda}{\partial\tilde\tau^2} = -\left(\frac{\partial l_j^\lambda}{\partial\tilde\tau}\right)^2 + \frac{\partial l_j^\lambda}{\partial\tilde\tau} + \frac{\tau^2}{L_j}\left(\frac{\pi_1 f_1}{\Phi\left(-\frac{t\sigma_{X_j}}{\sqrt{\eta^{-1}+\sigma^2_{X_j}}}\right)}\left(\left(\frac{\partial\log f_1}{\partial\tau}\right)^2 + \frac{\partial^2\log f_1}{\partial\tau^2}\right) + \right.$$

$$\left.\frac{\pi_3 f_3}{\Phi\left(-t\right)}\left(\left(\frac{\partial\log f_3}{\partial\tau}\right)^2 + \frac{\partial^2\log f_3}{\partial\tau^2}\right)\right)$$

$$\frac{\partial^2 l_j^\lambda}{\partial\tilde\tau\partial\tilde\eta} = -\frac{\partial l_j^\lambda}{\partial\tilde\tau}\frac{\partial l_j^\lambda}{\partial\tilde\eta} + \frac{\eta\tau\pi_1 f_1}{L_j\Phi\left(-\frac{t\sigma_{X_j}}{\sqrt{\eta^{-1}+\sigma^2_{X_j}}}\right)}\left(\frac{\partial\log f_1}{\partial\tau}\left(\frac{\partial\log f_1}{\partial\eta} - \frac{\partial\log\Phi\left(-\frac{t\sigma_{X_j}}{\sqrt{\eta^{-1}+\sigma^2_{X_j}}}\right)}{\partial\eta}\right) + \frac{\partial^2\log f_1}{\partial\tau\partial\eta}\right)$$

$$\frac{\partial^2 l_j^\lambda}{\partial\tilde{\tau}\partial\tilde{\pi}_1} = -\frac{\partial l_j^\lambda}{\partial\tilde{\tau}}\frac{\partial l_j^\lambda}{\partial\tilde{\pi}_1} + \frac{\tau}{L_j}\left(\frac{\pi_1(1-\pi_1)f_1}{\Phi\left(-\frac{t\sigma_{X_j}}{\sqrt{\eta^{-1}+\sigma_{X_j}^2}}\right)}\frac{\partial\log f_1}{\partial\tau} - \frac{\pi_3\pi_1 f_3}{\Phi(-t)}\frac{\partial\log f_3}{\partial\tau}\right)$$

$$\frac{\partial^2 l_j^\lambda}{\partial\tilde{\tau}\partial\tilde{\pi}_2} = -\frac{\partial l_j^\lambda}{\partial\tilde{\tau}}\frac{\partial l_j^\lambda}{\partial\tilde{\pi}_2} + \frac{\tau}{L_j}\left(\frac{-\pi_1\pi_2 f_1}{\Phi\left(-\frac{t\sigma_{X_j}}{\sqrt{\eta^{-1}+\sigma_{X_j}^2}}\right)}\frac{\partial\log f_1}{\partial\tau} - \frac{\pi_3\pi_2 f_3}{\Phi(-t)}\frac{\partial\log f_3}{\partial\tau}\right)$$

$$\frac{\partial^2 l_j^\lambda}{\partial\tilde{\tau}\partial\tilde{\pi}_3} = -\frac{\partial l_j^\lambda}{\partial\tilde{\tau}}\frac{\partial l_j^\lambda}{\partial\tilde{\pi}_3} + \frac{\tau}{L_j}\left(\frac{-\pi_1\pi_3 f_1}{\Phi\left(-\frac{t\sigma_{X_j}}{\sqrt{\eta^{-1}+\sigma_{X_j}^2}}\right)}\frac{\partial\log f_1}{\partial\tau} + \frac{\pi_3(1-\pi_3) f_3}{\Phi(-t)}\frac{\partial\log f_3}{\partial\tau}\right)$$

$$\frac{\partial^2 l_j^\lambda}{\partial\tilde{\eta}^2} = -\left(\frac{\partial l_j^\lambda}{\partial\tilde{\eta}}\right)^2 + \frac{\partial l_j^\lambda}{\partial\tilde{\eta}} + \frac{\eta^2}{L_j}\left(\frac{\pi_1 f_1}{\Phi\left(-\frac{t\sigma_{X_j}}{\sqrt{\eta^{-1}+\sigma_{X_j}^2}}\right)}\left(\left(\frac{\partial\log f_1}{\partial\eta} - \frac{\partial\log\Phi\left(-\frac{t\sigma_{X_j}}{\sqrt{\eta^{-1}+\sigma_{X_j}^2}}\right)}{\partial\eta}\right)^2 + \right.\right.$$

$$\left.\frac{\partial^2\log f_1}{\partial\eta^2} - \frac{\partial^2\log\Phi\left(-\frac{t\sigma_{X_j}}{\sqrt{\eta^{-1}+\sigma_{X_j}^2}}\right)}{\partial\eta^2}\right) + \frac{\pi_2 f_2}{\Phi\left(-\frac{t\sigma_{X_j}}{\sqrt{\eta^{-1}+\sigma_{X_j}^2}}\right)}\left(\left(\frac{\partial\log f_2}{\partial\eta} - \frac{\partial\log\Phi\left(-\frac{t\sigma_{X_j}}{\sqrt{\eta^{-1}+\sigma_{X_j}^2}}\right)}{\partial\eta}\right)^2 + \right.$$

$$\left.\left.\frac{\partial^2\log f_2}{\partial\eta^2} - \frac{\partial^2\log\Phi\left(-\frac{t\sigma_{X_j}}{\sqrt{\eta^{-1}+\sigma_{X_j}^2}}\right)}{\partial\eta^2}\right)\right)$$

$$\frac{\partial^2 l_j^\lambda}{\partial\tilde{\eta}\partial\tilde{\pi}_1} = -\frac{\partial l_j^\lambda}{\partial\tilde{\eta}}\frac{\partial l_j^\lambda}{\partial\tilde{\pi}_1} + \frac{\eta}{L_j}\left(\frac{\pi_1(1-\pi_1)f_1}{\Phi\left(-\frac{t\sigma_{X_j}}{\sqrt{\eta^{-1}+\sigma_{X_j}^2}}\right)}\left(\frac{\partial\log f_1}{\partial\eta} - \frac{\partial\log\Phi\left(-\frac{t\sigma_{X_j}}{\sqrt{\eta^{-1}+\sigma_{X_j}^2}}\right)}{\partial\eta}\right) - \right.$$

$$\left.\frac{\pi_2\pi_1 f_2}{\Phi\left(-\frac{t\sigma_{X_j}}{\sqrt{\eta^{-1}+\sigma_{X_j}^2}}\right)}\left(\frac{\partial\log f_2}{\partial\eta} - \frac{\partial\log\Phi\left(-\frac{t\sigma_{X_j}}{\sqrt{\eta^{-1}+\sigma_{X_j}^2}}\right)}{\partial\eta}\right)\right)$$

$$\frac{\partial^2 l_j^\lambda}{\partial\tilde{\eta}\partial\tilde{\pi}_2} = -\frac{\partial l_j^\lambda}{\partial\tilde{\eta}}\frac{\partial l_j^\lambda}{\partial\tilde{\pi}_2} + \frac{\eta}{L_j}\left(\frac{-\pi_1\pi_2 f_1}{\Phi\left(-\frac{t\sigma_{X_j}}{\sqrt{\eta^{-1}+\sigma^2_{X_j}}}\right)}\left(\frac{\partial\log f_1}{\partial\eta} - \frac{\partial\log\Phi\left(-\frac{t\sigma_{X_j}}{\sqrt{\eta^{-1}+\sigma^2_{X_j}}}\right)}{\partial\eta}\right) + \frac{\pi_2(1-\pi_2)f_2}{\Phi\left(-\frac{t\sigma_{X_j}}{\sqrt{\eta^{-1}+\sigma^2_{X_j}}}\right)}\left(\frac{\partial\log f_2}{\partial\eta} - \frac{\partial\log\Phi\left(-\frac{t\sigma_{X_j}}{\sqrt{\eta^{-1}+\sigma^2_{X_j}}}\right)}{\partial\eta}\right)\right)$$

$$\frac{\partial^2 l_j^\lambda}{\partial\tilde{\eta}\partial\tilde{\pi}_3} = -\frac{\partial l_j^\lambda}{\partial\tilde{\eta}}\frac{\partial l_j^\lambda}{\partial\tilde{\pi}_3} + \frac{\eta}{L_j}\left(\frac{-\pi_1\pi_3 f_1}{\Phi\left(-\frac{t\sigma_{X_j}}{\sqrt{\eta^{-1}+\sigma^2_{X_j}}}\right)}\left(\frac{\partial\log f_1}{\partial\eta} - \frac{\partial\log\Phi\left(-\frac{t\sigma_{X_j}}{\sqrt{\eta^{-1}+\sigma^2_{X_j}}}\right)}{\partial\eta}\right) - \frac{\pi_2\pi_3 f_2}{\Phi\left(-\frac{t\sigma_{X_j}}{\sqrt{\eta^{-1}+\sigma^2_{X_j}}}\right)}\left(\frac{\partial\log f_2}{\partial\eta} - \frac{\partial\log\Phi\left(-\frac{t\sigma_{X_j}}{\sqrt{\eta^{-1}+\sigma^2_{X_j}}}\right)}{\partial\eta}\right)\right)$$

$$\frac{\partial^2 l_j^\lambda}{\partial\tilde{\pi}_1^2} = -\left(\frac{\pi_1 f_1}{L_j\Phi\left(-\frac{t\sigma_{X_j}}{\sqrt{\eta^{-1}+\sigma^2_{X_j}}}\right)} - \pi_1\right)\left(\frac{\pi_1 f_1}{L_j\Phi\left(-\frac{t\sigma_{X_j}}{\sqrt{\eta^{-1}+\sigma^2_{X_j}}}\right)} + \pi_1\right) + \left(\frac{\pi_1 f_1}{L_j\Phi\left(-\frac{t\sigma_{X_j}}{\sqrt{\eta^{-1}+\sigma^2_{X_j}}}\right)} - \pi_1\right) - \frac{4\lambda\pi_1(1-\pi_1)}{m}$$

$$\frac{\partial^2 l_j^\lambda}{\partial\tilde{\pi}_1\partial\tilde{\pi}_2} = -\pi_1\pi_2\left(\frac{f_1 f_2}{L_j^2\Phi^2\left(-\frac{t\sigma_{X_j}}{\sqrt{\eta^{-1}+\sigma^2_{X_j}}}\right)} - 1 - \frac{4\lambda}{m}\right)$$

$$\frac{\partial^2 l_j^\lambda}{\partial\tilde{\pi}_1\partial\tilde{\pi}_3} = -\pi_1\pi_3\left(\frac{f_1 f_3}{L_j^2\Phi\left(-t\right)\Phi\left(-\frac{t\sigma_{X_j}}{\sqrt{\eta^{-1}+\sigma^2_{X_j}}}\right)} - 1 - \frac{4\lambda}{m}\right)$$

$$\frac{\partial^2 l_j^\lambda}{\partial\tilde{\pi}_2^2} = -\left(\frac{\pi_2 f_2}{L_j\Phi\left(-\frac{t\sigma_{X_j}}{\sqrt{\eta^{-1}+\sigma^2_{X_j}}}\right)} - \pi_2\right)\left(\frac{\pi_2 f_2}{L_j\Phi\left(-\frac{t\sigma_{X_j}}{\sqrt{\eta^{-1}+\sigma^2_{X_j}}}\right)} + \pi_2\right) + \left(\frac{\pi_2 f_2}{L_j\Phi\left(-\frac{t\sigma_{X_j}}{\sqrt{\eta^{-1}+\sigma^2_{X_j}}}\right)} - \pi_2\right)$$

$$
\begin{aligned}
& -\frac{4\lambda\pi_2(1-\pi_2)}{m} \\
\frac{\partial^2 l_j^\lambda}{\partial\tilde{\pi}_2\partial\tilde{\pi}_3} &= -\pi_2\pi_3\left(\frac{f_2 f_3}{L_j^2\Phi\left(-t\right)\Phi\left(-\frac{t\sigma_{X_j}}{\sqrt{\eta^{-1}+\sigma_{X_j}^2}}\right)}-1-\frac{4\lambda}{m}\right) \\
\frac{\partial^2 l_j^\lambda}{\partial\tilde{\pi}_3^2} &= -\left(\frac{\pi_3 f_3}{L_j\Phi\left(-t\right)}-\pi_3\right)\left(\frac{\pi_3 f_3}{L_j\Phi\left(-t\right)}+\pi_3\right)+\left(\frac{\pi_3 f_3}{L_j\Phi\left(-t\right)}-\pi_3\right)-\frac{4\lambda\pi_3(1-\pi_3)}{m}.
\end{aligned}
\tag{S2}
$$

We omit the "2" preceding $\Phi$ because it can be factored out as $-\log 2$ in the log-likelihood function. Since this constant term does not influence the optimization or the final results, it is excluded for simplicity. The $f_1, f_2, f_3, f_4$ are the normal density functions of the four components, and $L_j$ is the likelihood function of $\hat{d}_j$, i.e., $\frac{\pi_1 f_1}{\Phi\left(-\frac{t\sigma_{X_j}}{\sqrt{\eta^{-1}+\sigma_{X_j}^2}}\right)}+\frac{\pi_2 f_2}{\Phi\left(-\frac{t\sigma_{X_j}}{\sqrt{\eta^{-1}+\sigma_{X_j}^2}}\right)}+\frac{\pi_3 f_3}{\Phi(-t)}+\frac{\pi_4 f_4}{\Phi(-t)}$. Then the Sandwitch estimator of the variance of $\hat{\beta}$ is obtained by replacing the expectations with the sample averages.

#### 1.2 Details of DAEM algorithm

The complete data penalized log-likelihood function of the GMM model is given by

$$
\begin{aligned}
& l_{c,j}^\lambda(\theta;\alpha_{j0},\gamma_{j0},L_j,\hat{\Gamma}_j,\hat{\gamma}_j,\sigma_{Y_j},\sigma_{X_j}) \\
=& L_{j1}\left(\log\pi_1 N\left(\hat{d}_j;\begin{bmatrix}\alpha_{j0}+\beta\gamma_{j0}\\ \gamma_{j0}\end{bmatrix},\begin{bmatrix}\sigma_{Y_j}^2 & 0\\ 0 & \sigma_{X_j}^2\end{bmatrix}\right)N\left(\alpha_{j0};0,\tau^{-1}\right)N\left(\gamma_{j0};0,\eta^{-1}\right)-\log\Phi\left(-\frac{t\sigma_{X_j}}{\sqrt{\eta^{-1}+\sigma_{X_j}^2}}\right)\right)+ \\
& L_{j2}\left(\log\pi_2 N\left(\hat{d}_j;\begin{bmatrix}\beta\gamma_{j0}\\ \gamma_{j0}\end{bmatrix},\begin{bmatrix}\sigma_{Y_j}^2 & 0\\ 0 & \sigma_{X_j}^2\end{bmatrix}\right)N\left(\alpha_{j0};0,\tau^{-1}\right)N\left(\gamma_{j0};0,\eta^{-1}\right)-\log\Phi\left(-\frac{t\sigma_{X_j}}{\sqrt{\eta^{-1}+\sigma_{X_j}^2}}\right)\right)+ \\
& L_{j3}\left(\log\pi_3 N\left(\hat{d}_j;\begin{bmatrix}\alpha_{j0}\\ 0\end{bmatrix},\begin{bmatrix}\sigma_{Y_j}^2 & 0\\ 0 & \sigma_{X_j}^2\end{bmatrix}\right)N\left(\alpha_{j0};0,\tau^{-1}\right)N\left(\gamma_{j0};0,\eta^{-1}\right)-\log\Phi\left(-t\right)\right)+ \\
& L_{j4}\left(\log\pi_4 N\left(\hat{d}_j;\begin{bmatrix}0\\ 0\end{bmatrix},\begin{bmatrix}\sigma_{Y_j}^2 & 0\\ 0 & \sigma_{X_j}^2\end{bmatrix}\right)N\left(\alpha_{j0};0,\tau^{-1}\right)N\left(\gamma_{j0};0,\eta^{-1}\right)-\log\Phi\left(-t\right)\right)-(\sum_{k=1}^{4}\pi_k-1)+\lambda\sum_{k=1}^{4}\log\pi_k,
\end{aligned}
\tag{S3}
$$

For E-step, the posterior probabilities of the latent variables are derived based on Equation (S3). After factoring the joint posterior probabilities as

$$
\begin{aligned}
&p(\alpha_{j0}, \gamma_{j0}, L_j|\hat{\Gamma}_j, \hat{\gamma}_j, \sigma_{Y_j}, \sigma_{X_j}) \\
=&(p(\alpha_{j0}, \gamma_{j0}|L_{j1} = 1, \hat{\Gamma}_j, \hat{\gamma}_j, \sigma_{Y_j}, \sigma_{X_j})p(L_{j1} = 1|\hat{\Gamma}_j, \hat{\gamma}_j, \sigma_{Y_j}, \sigma_{X_j}))^{L_{j1}} \\
&(p(\alpha_{j0}, \gamma_{j0}|L_{j2} = 1, \hat{\Gamma}_j, \hat{\gamma}_j, \sigma_{Y_j}, \sigma_{X_j})p(L_{j2} = 1|\hat{\Gamma}_j, \hat{\gamma}_j, \sigma_{Y_j}, \sigma_{X_j}))^{L_{j2}} \\
&(p(\alpha_{j0}, \gamma_{j0}|L_{j3} = 1, \hat{\Gamma}_j, \hat{\gamma}_j, \sigma_{Y_j}, \sigma_{X_j})p(L_{j3} = 1|\hat{\Gamma}_j, \hat{\gamma}_j, \sigma_{Y_j}, \sigma_{X_j}))^{L_{j3}} \\
&(p(\alpha_{j0}, \gamma_{j0}|L_{j4} = 1, \hat{\Gamma}_j, \hat{\gamma}_j, \sigma_{Y_j}, \sigma_{X_j})p(L_{j4} = 1|\hat{\Gamma}_j, \hat{\gamma}_j, \sigma_{Y_j}, \sigma_{X_j}))^{L_{j4}},
\end{aligned}
\tag{S4}
$$

one can obtain the posterior probabilities of the latent variables as

$$
\begin{aligned}
p(\alpha_{j0}, \gamma_{j0}|L_{jk} = 1, \hat{\Gamma}_j, \hat{\gamma}_j, \sigma_{Y_j}, \sigma_{X_j}) =& N(\mu_{jk}, \Sigma_{jk}), \\
p(L_{jk} = 1|\hat{\Gamma}_j, \hat{\gamma}_j, \sigma_{Y_j}, \sigma_{X_j}) =& \rho_{jk},
\end{aligned}
\tag{S5}
$$

for $k = 1, 2, 3, 4$, where

$$
\begin{aligned}
\mu_{j1} = \Sigma_{j1}\begin{bmatrix} \hat{\Gamma}_j\sigma_{Y_j}^{-2} \\ \beta\hat{\Gamma}_j\sigma_{Y_j}^{-2} + \hat{\gamma}_j\sigma_{X_j}^{-2} \end{bmatrix}, \Sigma_{j1} &= \begin{bmatrix} \tau + \sigma_{Y_j}^{-2} & \beta\sigma_{Y_j}^{-2} \\ \beta\sigma_{Y_j}^{-2} & \eta + \beta^2\sigma_{Y_j}^{-2} + \sigma_{X_j}^{-2} \end{bmatrix}^{-1}, \\
\mu_{j2} = \Sigma_{j2}\begin{bmatrix} 0 \\ \beta\hat{\Gamma}_j\sigma_{Y_j}^{-2} + \hat{\gamma}_j\sigma_{X_j}^{-2} \end{bmatrix}, \Sigma_{j2} &= \begin{bmatrix} \tau & 0 \\ 0 & \eta + \beta^2\sigma_{Y_j}^{-2} + \sigma_{X_j}^{-2} \end{bmatrix}^{-1}, \\
\mu_{j3} = \Sigma_{j3}\begin{bmatrix} \hat{\Gamma}_j\sigma_{Y_j}^{-2} \\ 0 \end{bmatrix}, \Sigma_{j3} &= \begin{bmatrix} \tau + \sigma_{Y_j}^{-2} & 0 \\ 0 & \eta \end{bmatrix}^{-1}, \\
\mu_{j4} = \Sigma_{j4}\begin{bmatrix} 0 \\ 0 \end{bmatrix}, \Sigma_{j4} &= \begin{bmatrix} \tau & 0 \\ 0 & \eta \end{bmatrix}^{-1},
\end{aligned}
\tag{S6}
$$

and

$$
\begin{aligned}
\rho_{jk} =& \frac{\pi_k \exp\{\frac{1}{2}\log|\Sigma_{jk}| + \frac{1}{2}\mu_{jk}^T\Sigma_{jk}^{-1}\mu_{jk}\}/\Phi_k}{\sum_{k=1}^4 \pi_k \exp\{\frac{1}{2}\log|\Sigma_{jk}| + \frac{1}{2}\mu_{jk}^T\Sigma_{jk}^{-1}\mu_{jk}\}/\Phi_k}, \\
\Phi_k =& \begin{cases} \Phi\left(-\frac{t\sigma_{X_j}}{\sqrt{\eta^{-1}+\sigma_{X_j}^2}}\right) & \text{for k=1, 2} \\ \Phi(-t) & \text{for k=3, 4} \end{cases}.
\end{aligned}
\tag{S7}
$$

For M-step, the parameters are updated by maximizing the expected complete data penalized log-likelihood function. The update of the parameters is given by

$$
\begin{aligned}
\hat{\beta} =& \frac{\sum_{j=1}^{m} \rho_{j1}\sigma_{Y_j}^{-2}\left(\mu_{j1,2}(\hat{\Gamma}_j - \mu_{j1,1}) - \Sigma_{j1,12}\right) + \rho_{j2}\sigma_{Y_j}^{-2}\mu_{j2,2}\hat{\Gamma}_j}{\sum_{j=1}^{m} \rho_{j1}\sigma_{Y_j}^{-2}\left(\mu_{j1,2}^2 + \Sigma_{j1,22}^2\right) + \rho_{j2}\sigma_{Y_j}^{-2}\left(\mu_{j2,2}^2 + \Sigma_{j2,22}^2\right)}, \\
\hat{\tau} =& m / \sum_{j=1}^{m}\sum_{k=1}^{4} \rho_{jk}\left(\mu_{jk,1}^2 + \Sigma_{jk,11}^2\right), \qquad \text{(S8)} \\
\hat{\pi}_k =& (\lambda + \sum_{j=1}^{m} \rho_{jk})/(4\lambda + m), \text{ for } k = 1, 2, 3, 4,
\end{aligned}
$$

where $\mu_{jk,a}$ and $\Sigma_{jk,ab}$ represent the $a$th element and $(a,b)$th element of $\mu_{jk}$ and $\Sigma_{jk}$, respectively. However, $\hat{\eta}$ has no explicit form. It is the solution of the following equation:

$$
m/\eta + \sum_{j=1}^{m} \frac{\phi\left(-\frac{t\sigma_{X_j}}{\sqrt{\eta^{-1}+\sigma_{X_j}^2}}\right)}{\Phi\left(-\frac{t\sigma_{X_j}}{\sqrt{\eta^{-1}+\sigma_{X_j}^2}}\right)}(\rho_{j1}+\rho_{j2})t\sigma_{X_j}(\eta^{-1}+\sigma_{X_j}^2)^{-3/2}\eta^{-2} = \sum_{j=1}^{m}\sum_{k=1}^{4}\rho_{jk}\left(\mu_{jk,2}^2 + \Sigma_{jk,22}^2\right). \qquad \text{(S9)}
$$

We use the following fixed point iteration to update $\hat{\eta}$:

$$
\eta^{t+1} = \left( \frac{m/\sqrt{\eta^t} + \sum_{j=1}^{m} \frac{\phi\left(-\frac{t\sigma_{X_j}}{\sqrt{\eta^{-1}+\sigma_{X_j}^2}}\right)}{\Phi\left(-\frac{t\sigma_{X_j}}{\sqrt{\eta^{-1}+\sigma_{X_j}^2}}\right)}(\rho_{j1}+\rho_{j2})t\sigma_{X_j}(1+\sigma_{X_j}^2\eta^t)^{-3/2}}{\sum_{j=1}^{m}\sum_{k=1}^{4}\rho_{jk}\left(\mu_{jk,2}^2 + \Sigma_{jk,22}^2\right)} \right)^2. \qquad \text{(S10)}
$$

In summary, the DAEM algorithm is given as follows,

**Algorithm 1:** DAEM algorithm for the GMM model.

**Data:** $\hat{\Gamma}_j, \hat{\gamma}_j, \sigma_{Y_j}, \sigma_{X_j}$, for $j = 1, ..., m$
**Init:** $\theta^0 = (\beta^0, \tau^0, \eta^0, \pi_k^0)^T, 0 < \xi^0 < 1, ELBO^0$
**Const:** $\lambda \geq 1, r > 1, \varepsilon > 0$
**for** $t = 0, ..., \mathcal{T} - 1$ **do**
  Update $\mu_{jk}^t, \Sigma_{jk}^t, \rho_{jk}^t$ according to (S6) and (S7); `/* E-step */`
  $\rho_{jk}^t \leftarrow {\rho_{jk}^t}^{\xi^t} / \sum_{k=1}^4 {\rho_{jk}^t}^{\xi^t}$; `/* Annealing */`
  Update $\theta^{t+1}$ according to (7); `/* M-step */`
  $ELBO^{t+1} \leftarrow ELBO(\theta^t; \hat{\Gamma}, \hat{\gamma}, \sigma_Y, \sigma_X)$;
  $\xi^{t+1} \leftarrow \min(1, r\xi^t)$;
  **if** $|ELBO^{t+1} - ELBO^t| < \varepsilon$ **then**
    break;
  **end**
**end**

Computing the original penalized log-likelihood is time-consuming, so we use the evidence lower bound (ELBO) as the convergence criterion. The ELBO is defined as the expected complete data penalized log-likelihood function minus the entropy of the posterior distribution of the latent variables, which is given by

$$ELBO = \sum_{j=1}^{m} \sum_{k=1}^{4} \rho_{jk} \left( \frac{1}{2}(\log |\Sigma_{jk}| + \mu_{jk}^T \Sigma_{jk}^{-1} \mu_{jk}) + \log \pi_k - \log \rho_{jk} - \log \Phi_k \right) + \frac{1}{2}(\log \tau + \log \eta) + \lambda \sum_{k=1}^{4} \pi_k + const. \tag{S11}$$

### 2 Supplementary Figures

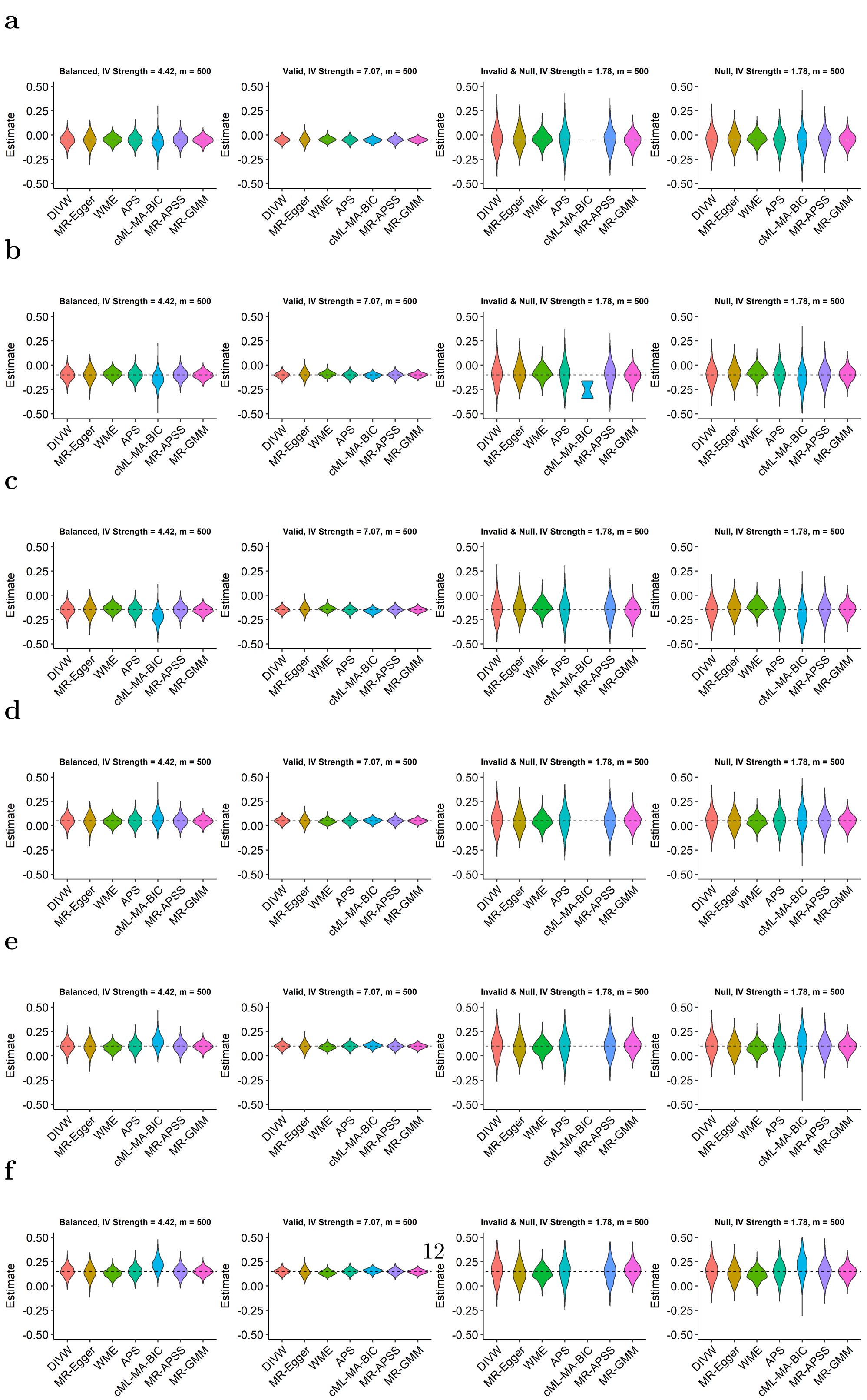


**Fig. S1**: **Comparison of point estimates under different $\beta$ values.** Violin plots display the distribution of point estimates from 1000 simulations for varying true causal effects, $\beta$. The dashed line in each plot indicates the true value of $\beta$. **a.** $\beta = -0.05$. **b.** $\beta = -0.10$. **c.** $\beta = -0.15$. **d.** $\beta = 0.05$. **e.** $\beta = 0.10$. **f.** $\beta = 0.15$.

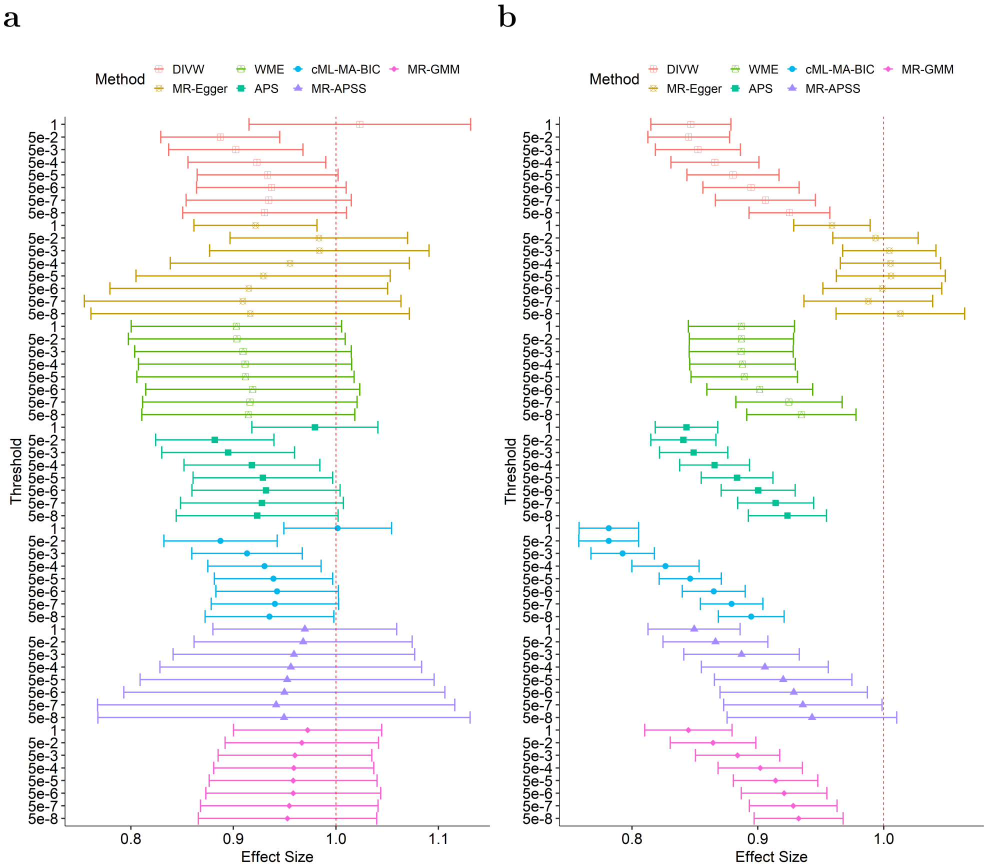


**Fig. S2**: **Benchmark HDL-HDL identity causal effect estimates obtained by using pseudo-$p$-value-based versus $p$-value-based LD clumping procedures under different IV selection thresholds. a.** The forest plot of all methods using pseudo-$p$-value-based LD clumping procedure, which is conducted by first clumping based on pseudo $p$-values and then thresholding based on observed $p$-values. The 95% confidence intervals of point estimates are represented by error bars. **b.** The forest plot of all methods using standard $p$-value-based LD clumping procedure. These plots allow for a visual comparison of how different clumping methods and IV selection thresholds affect the estimation of causal effects in MR studies.

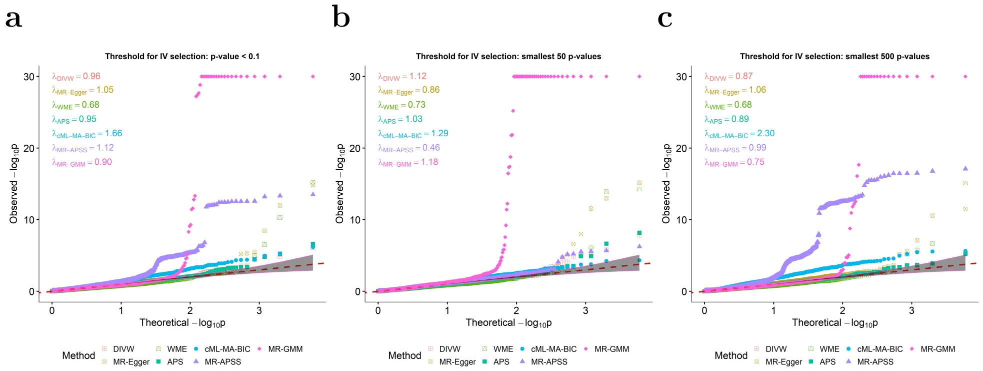


**Fig. S3**: **The Q-Q plot diagnosis of competing methods in the proteome-wide MR study under different $p$-value thresholds.** Q-Q plots of the negative log-transformed $p$-values for each method are presented, with the $\lambda_{\text{GC}}$ values listed in the top-left corner of each plot for quantitative comparison. The grey regions on the plots represent the 95% confidence intervals for these $p$-values. After the pseudo-$p$-value-based LD clumping procedure, SNPs are selected based on different $p$-value thresholds: **a.** threshold of $p$-value $< 0.1$; **b.** smallest 50 $p$-values **c.** smallest 500 $p$-values.

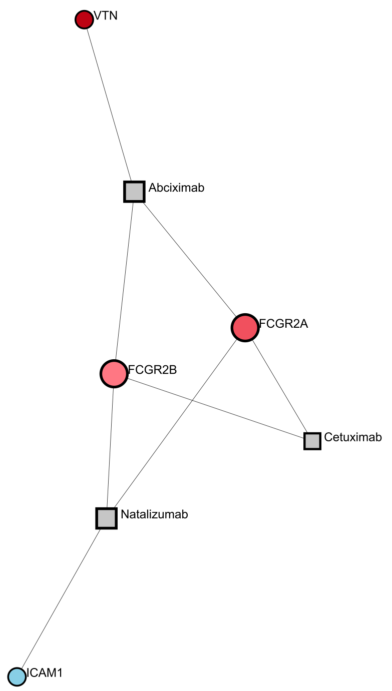


**Fig. S4**: The trimmed protein-drug interaction network of the 45 identified plasma proteins, with protein sizes proportional to their degrees. Proteins are color-coded according to their causal effects on CHD: red represents proteins that increase CHD risk, and blue indicates those that decrease it.

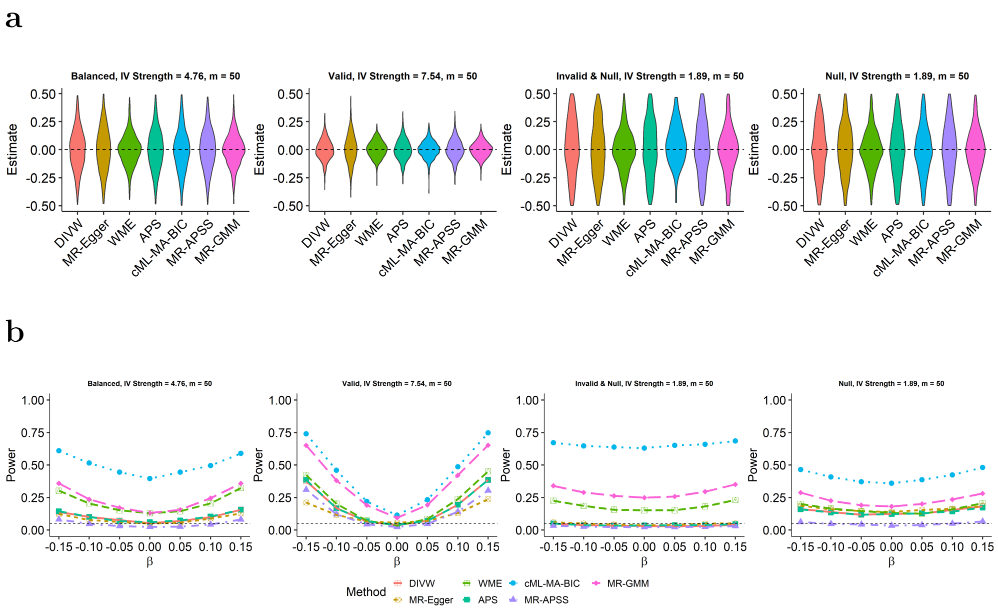


**Fig. S5**: **Benchmark all seven methods in various simulation scenarios when the number of IVs $m = 50$. a.** Violin plots depict the point estimates from 1000 replications when the true causal effect $\beta = 0$, indicated by the dashed line. **b.** The power of compared methods, calculated as the proportion of simulations in which the null hypothesis $H_0 : \beta = 0$ is rejected across 1000 replications.

**a**

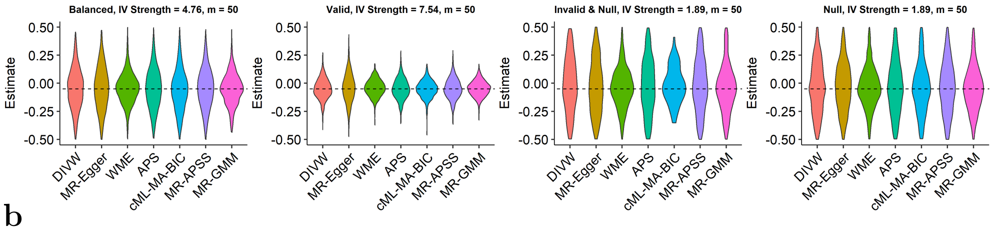


**b**

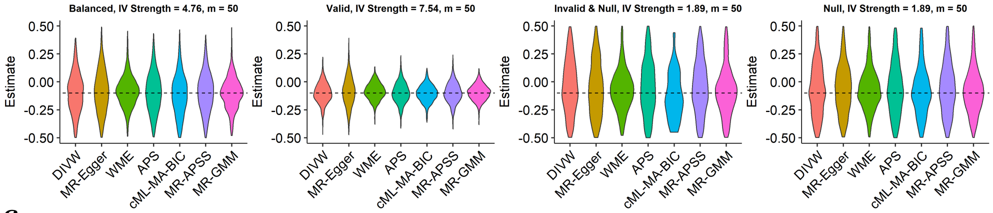


**c**

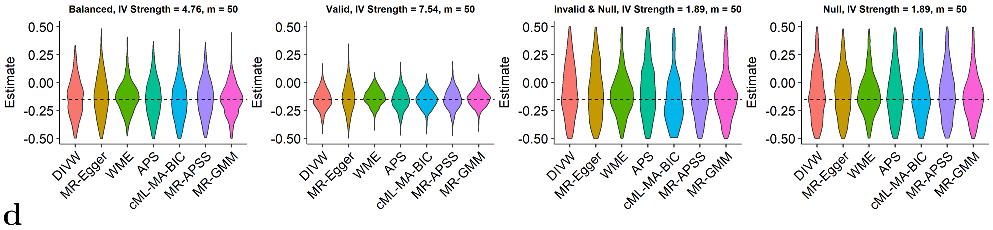


**d**

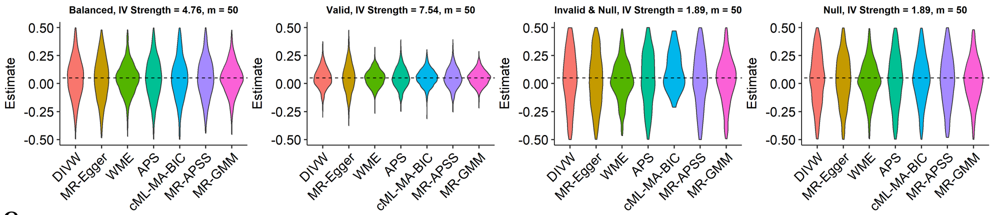


**e**

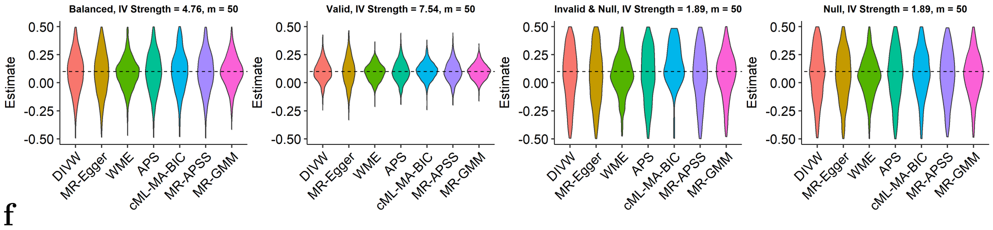


**f**

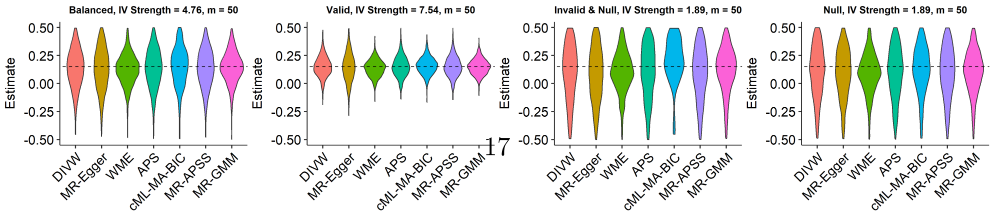




**Fig. S6**: **Comparison of point estimates under different $\beta$ values when the number of IVs $m = 50$.** Violin plots display the distribution of point estimates from 1000 simulations for varying true causal effects, $\beta$. The dashed line in each plot indicates the true value of $\beta$. **a.** $\beta = -0.05$. **b.** $\beta = -0.10$. **c.** $\beta = -0.15$. **d.** $\beta = 0.05$. **e.** $\beta = 0.10$. **f.** $\beta = 0.15$.

### 3 Supplementary Tables

| P | # of IVs | IV strength | Exposure ID | Outcome ID | DIVW | MR-Egger | WME | APS | cML-MA-BIC | MR-APSS | MR-GMM |
|---|---|---|---|---|---|---|---|---|---|---|---|
| 5e-08 | 46 | 71.669 | ukb-b-19953 | ieu-a-2 | 0.963(0.031) | 1.044(0.072) | 1.003(0.048) | 0.962(0.031) | 0.961(0.031) | 0.982(0.083) | 0.987(0.032) |
| 5e-07 | 60 | 61.143 | ukb-b-19953 | ieu-a-2 | 0.947(0.031) | 1.064(0.070) | 0.997(0.047) | 0.945(0.031) | 0.946(0.030) | 0.970(0.075) | 0.984(0.031) |
| 5e-06 | 76 | 52.901 | ukb-b-19953 | ieu-a-2 | **0.935(0.030)** | 1.072(0.066) | 0.994(0.045) | **0.934(0.030)** | **0.933(0.029)** | 0.961(0.068) | 0.977(0.030) |
| 5e-05 | 107 | 42.632 | ukb-b-19953 | ieu-a-2 | **0.929(0.029)** | 1.063(0.063) | 0.994(0.044) | **0.927(0.029)** | **0.927(0.028)** | 0.970(0.061) | 0.981(0.028) |
| 5e-04 | 162 | 32.636 | ukb-b-19953 | ieu-a-2 | **0.916(0.027)** | 1.049(0.057) | 0.980(0.041) | **0.913(0.027)** | **0.914(0.027)** | 0.967(0.054) | 0.983(0.027) |
| 0.005 | 253 | 24.015 | ukb-b-19953 | ieu-a-2 | **0.903(0.025)** | 1.047(0.050) | 0.925(0.040) | **0.899(0.025)** | **0.895(0.027)** | 0.961(0.047) | 0.976(0.027) |
| 0.050 | 479 | 14.856 | ukb-b-19953 | ieu-a-2 | **0.878(0.022)** | 1.047(0.039) | **0.880(0.038)** | **0.875(0.022)** | **0.854(0.024)** | 0.950(0.040) | 0.962(0.026) |
| 1.000 | 1773 | 4.048 | ukb-b-19953 | ieu-a-2 | 0.961(0.022) | **0.886(0.024)** | **0.879(0.037)** | **0.932(0.024)** | **0.902(0.024)** | **0.934(0.032)** | **0.946(0.026)** |

**Table S1**: BMI-BMI causal effect estimates and SEs using the pseudo-$p$-value-based LD clumping procedure. Estimates with 95% confidence intervals that fail to include the true causal effect, $\beta = 1$, are highlighted in bold.

| P | # of IVs | IV strength | Exposure ID | Outcome ID | DIVW | MR-Egger | WME | APS | cML-MA-BIC | MR-APSS | MR-GMM |
|---|---|---|---|---|---|---|---|---|---|---|---|
| 5e-08 | 405 | 63.031 | ukb-b-19953 | ieu-a-2 | **0.841(0.015)** | **1.096(0.034)** | **0.864(0.021)** | **0.830(0.015)** | **0.793(0.015)** | **0.859(0.029)** | **0.855(0.018)** |
| 5e-07 | 510 | 55.505 | ukb-b-19953 | ieu-a-2 | **0.827(0.013)** | **1.072(0.030)** | **0.853(0.020)** | **0.816(0.014)** | **0.771(0.016)** | **0.850(0.026)** | **0.844(0.017)** |
| 5e-06 | 620 | 49.566 | ukb-b-19953 | ieu-a-2 | **0.812(0.013)** | **1.066(0.029)** | **0.839(0.019)** | **0.801(0.013)** | **0.751(0.013)** | **0.836(0.024)** | **0.834(0.017)** |
| 5e-05 | 774 | 43.193 | ukb-b-19953 | ieu-a-2 | **0.802(0.012)** | 1.045(0.026) | **0.824(0.019)** | **0.792(0.012)** | **0.741(0.012)** | **0.822(0.022)** | **0.821(0.016)** |
| 5e-04 | 940 | 37.873 | ukb-b-19953 | ieu-a-2 | **0.794(0.011)** | 1.021(0.024) | **0.815(0.018)** | **0.785(0.011)** | **0.731(0.011)** | **0.813(0.020)** | **0.811(0.015)** |
| 0.005 | 1156 | 32.433 | ukb-b-19953 | ieu-a-2 | **0.783(0.011)** | 1.009(0.022) | **0.807(0.018)** | **0.773(0.011)** | **0.719(0.011)** | **0.800(0.019)** | **0.800(0.015)** |
| 0.050 | 1383 | 27.907 | ukb-b-19953 | ieu-a-2 | **0.776(0.011)** | 0.985(0.020) | **0.798(0.017)** | **0.767(0.011)** | **0.712(0.011)** | **0.782(0.017)** | **0.783(0.015)** |
| 1.000 | 1820 | 21.326 | ukb-b-19953 | ieu-a-2 | **0.780(0.011)** | **0.911(0.017)** | **0.797(0.017)** | **0.772(0.010)** | **0.709(0.011)** | **0.770(0.016)** | **0.768(0.015)** |

**Table S2**: BMI-BMI causal effect estimates and SEs using the classic $p$-value-based LD clumping procedure. Estimates with 95% confidence intervals that fail to include the true causal effect, $\beta = 1$, are highlighted in bold.

| P | # of IVs | IV strength | Exposure ID | Outcome ID | DIVW | MR-Egger | WME | APS | cML-MA-BIC | MR-APSS | MR-GMM |
|---|---|---|---|---|---|---|---|---|---|---|---|
| 5e-08 | 40 | 105.400 | ieu-b-109 | ebi-a-GCST002223 | 0.930(0.041) | 0.916(0.079) | 0.915(0.053) | 0.923(0.040) | **0.935(0.032)** | 0.949(0.093) | 0.953(0.044) |
| 5e-07 | 44 | 98.277 | ieu-b-109 | ebi-a-GCST002223 | 0.934(0.041) | 0.909(0.079) | 0.916(0.053) | 0.928(0.041) | 0.940(0.032) | 0.942(0.089) | 0.954(0.044) |
| 5e-06 | 59 | 78.905 | ieu-b-109 | ebi-a-GCST002223 | 0.937(0.037) | 0.915(0.069) | 0.919(0.053) | 0.932(0.037) | 0.943(0.030) | 0.950(0.080) | 0.958(0.043) |
| 5e-05 | 78 | 63.886 | ieu-b-109 | ebi-a-GCST002223 | 0.933(0.035) | 0.929(0.063) | 0.912(0.054) | **0.929(0.035)** | **0.939(0.029)** | 0.952(0.073) | 0.958(0.042) |
| 5e-04 | 113 | 48.175 | ieu-b-109 | ebi-a-GCST002223 | **0.923(0.034)** | 0.955(0.059) | 0.911(0.053) | **0.918(0.034)** | **0.930(0.028)** | 0.956(0.065) | 0.959(0.040) |
| 0.005 | 178 | 33.723 | ieu-b-109 | ebi-a-GCST002223 | **0.902(0.033)** | 0.984(0.055) | 0.909(0.054) | **0.895(0.033)** | **0.913(0.028)** | 0.959(0.060) | 0.960(0.038) |
| 0.050 | 389 | 17.863 | ieu-b-109 | ebi-a-GCST002223 | **0.887(0.030)** | 0.983(0.044) | 0.903(0.054) | **0.882(0.029)** | **0.887(0.028)** | 0.968(0.054) | 0.967(0.038) |
| 1.000 | 1775 | 3.850 | ieu-b-109 | ebi-a-GCST002223 | 1.023(0.055) | **0.921(0.031)** | 0.903(0.052) | 0.979(0.031) | 1.002(0.027) | 0.970(0.046) | 0.972(0.037) |

**Table S3**: HDL-HDL causal effect estimates and SEs using the pseudo-$p$-value-based LD clumping procedure. Estimates with 95% confidence intervals that fail to include the true causal effect, $\beta = 1$, are highlighted in bold.

| P | # of IVs | IV strength | Exposure ID | Outcome ID | DIVW | MR-Egger | WME | APS | cML-MA-BIC | MR-APSS | MR-GMM |
|---|---|---|---|---|---|---|---|---|---|---|---|
| 5e-08 | 313 | 127.690 | ieu-b-109 | ebi-a-GCST002223 | **0.925(0.016)** | 1.013(0.026) | **0.935(0.022)** | **0.924(0.016)** | **0.895(0.013)** | 0.943(0.034) | **0.932(0.018)** |
| 5e-07 | 369 | 112.260 | ieu-b-109 | ebi-a-GCST002223 | **0.906(0.020)** | 0.988(0.026) | **0.925(0.022)** | **0.914(0.015)** | **0.879(0.013)** | **0.936(0.032)** | **0.928(0.018)** |
| 5e-06 | 461 | 94.237 | ieu-b-109 | ebi-a-GCST002223 | **0.895(0.020)** | 0.999(0.024) | **0.902(0.021)** | **0.900(0.015)** | **0.865(0.013)** | **0.928(0.030)** | **0.921(0.017)** |
| 5e-05 | 592 | 77.234 | ieu-b-109 | ebi-a-GCST002223 | **0.880(0.019)** | 1.006(0.022) | **0.889(0.022)** | **0.884(0.015)** | **0.846(0.013)** | **0.920(0.028)** | **0.914(0.017)** |
| 5e-04 | 775 | 62.090 | ieu-b-109 | ebi-a-GCST002223 | **0.866(0.018)** | 1.005(0.020) | **0.888(0.021)** | **0.866(0.014)** | **0.827(0.014)** | **0.906(0.026)** | **0.902(0.017)** |
| 0.005 | 1021 | 49.242 | ieu-b-109 | ebi-a-GCST002223 | **0.852(0.017)** | 1.004(0.019) | **0.887(0.021)** | **0.849(0.014)** | **0.793(0.013)** | **0.887(0.023)** | **0.884(0.017)** |
| 0.050 | 1310 | 39.408 | ieu-b-109 | ebi-a-GCST002223 | **0.845(0.017)** | 0.993(0.017) | **0.887(0.021)** | **0.841(0.013)** | **0.782(0.012)** | **0.866(0.021)** | **0.864(0.017)** |
| 1.000 | 1809 | 28.678 | ieu-b-109 | ebi-a-GCST002223 | **0.847(0.016)** | **0.959(0.015)** | **0.887(0.022)** | **0.843(0.013)** | **0.782(0.012)** | **0.849(0.019)** | **0.845(0.018)** |

**Table S4**: HDL-HDL causal effect estimates and SEs using the classic $p$-value-based LD clumping procedure. Estimates with 95% confidence intervals that fail to include the true causal effect, $\beta = 1$, are highlighted in bold.

| Exposure ID | Exposure | Outcome ID | Outcome | \# of IVs | IV strength | Estimate | SE | P-value |
|---|---|---|---|---|---|---|---|---|
| ieu-a-1024 | Multiple sclerosis | ukb-d-1747_1 | hair color | 97 | 7.455 | -0.001 | 0.005 | 0.826 |
| ieu-a-789 | Urate | ukb-d-1747_1 | hair color | 88 | 5.113 | -0.001 | 0.011 | 0.941 |
| ieu-a-1097 | Microalbuminuria | ukb-d-1747_1 | hair color | 81 | 4.648 | 0.012 | 0.017 | 0.485 |
| ieu-a-1049 | Iron | ukb-d-1747_1 | hair color | 107 | 4.697 | -0.017 | 0.034 | 0.627 |
| ieu-a-961 | Cigarettes smoked per day | ukb-d-1747_1 | hair color | 92 | 4.743 | 0.001 | 0.004 | 0.740 |
| ieu-a-1085 | Amyotrophic lateral sclerosis | ukb-d-1747_1 | hair color | 138 | 4.758 | 0.065 | 0.083 | 0.428 |
| ieu-a-964 | Age of smoking initiation | ukb-d-1747_1 | hair color | 94 | 4.850 | 0.157 | 0.132 | 0.235 |
| ieu-a-801 | Bipolar disorder | ukb-d-1747_1 | hair color | 136 | 4.821 | 0.003 | 0.016 | 0.876 |
| ieu-a-1170 | Knee and hip osteoarthritis | ukb-d-1747_1 | hair color | 95 | 4.765 | -0.005 | 0.014 | 0.711 |
| ieu-a-989 | Squamous cell lung cancer | ukb-d-1747_1 | hair color | 161 | 5.497 | -0.006 | 0.004 | 0.149 |
| ieu-a-1052 | Transferrin | ukb-d-1747_1 | hair color | 134 | 5.062 | 0.006 | 0.011 | 0.585 |
| ieu-a-55 | Hip circumference | ukb-d-1747_1 | hair color | 170 | 9.228 | 0.011 | 0.009 | 0.225 |
| ieu-a-12 | Crohn's disease | ukb-d-1747_1 | hair color | 203 | 17.974 | 0.003 | 0.002 | 0.155 |
| ieu-a-91 | Obesity class 2 | ukb-d-1747_1 | hair color | 91 | 4.826 | -0.002 | 0.009 | 0.804 |
| ieu-a-27 | Birth weight | ukb-d-1747_1 | hair color | 119 | 4.617 | 0.004 | 0.062 | 0.945 |
| ieu-a-1044 | Caudate volume | ukb-d-1747_1 | hair color | 112 | 4.372 | 0.000 | 0.000 | 0.967 |
| ieu-a-835 | Body mass index | ukb-d-1747_1 | hair color | 203 | 10.075 | -0.006 | 0.009 | 0.516 |
| ieu-a-970 | Ulcerative colitis | ukb-d-1747_1 | hair color | 174 | 13.223 | -0.001 | 0.002 | 0.822 |
| ieu-a-85 | Extreme body mass index | ukb-d-1747_1 | hair color | 120 | 4.596 | -0.011 | 0.023 | 0.646 |
| ieu-a-28 | Infant head circumference | ukb-d-1747_1 | hair color | 97 | 5.071 | -0.010 | 0.010 | 0.313 |
| ieu-a-806 | Autism | ukb-d-1747_1 | hair color | 123 | 4.892 | 0.011 | 0.009 | 0.216 |
| ieu-a-1047 | Putamen volume | ukb-d-1747_1 | hair color | 124 | 4.623 | 0.000 | 0.000 | 0.789 |
| ieu-a-1041 | Intracranial volume | ukb-d-1747_1 | hair color | 110 | 4.865 | 0.000 | 0.000 | 0.723 |
| ieu-a-79 | Waist-to-hip ratio | ukb-d-1747_1 | hair color | 140 | 7.476 | 0.008 | 0.010 | 0.429 |
| ieu-a-803 | PGC cross-disorder traits | ukb-d-1747_1 | hair color | 135 | 5.124 | -0.004 | 0.011 | 0.721 |
| ieu-a-1169 | Hip osteoarthritis | ukb-d-1747_1 | hair color | 95 | 4.788 | -0.009 | 0.014 | 0.527 |
| ieu-a-1096 | Childhood obesity | ukb-d-1747_1 | hair color | 99 | 4.663 | -0.116 | 0.264 | 0.661 |
| ieu-a-276 | Celiac disease | ukb-d-1747_1 | hair color | 117 | 4.661 | 0.001 | 0.015 | 0.927 |
| ieu-a-984 | Lung adenocarcinoma | ukb-d-1747_1 | hair color | 136 | 5.054 | -0.009 | 0.007 | 0.157 |
| ieu-a-16 | Childhood intelligence | ukb-d-1747_1 | hair color | 105 | 4.808 | 0.003 | 0.016 | 0.853 |
| ieu-a-114 | Neo-conscientiousness | ukb-d-1747_1 | hair color | 109 | 4.463 | -0.051 | 0.061 | 0.399 |
| ieu-a-86 | Extreme height | ukb-d-1747_1 | hair color | 93 | 5.982 | 0.007 | 0.003 | 0.055 |
| ieu-a-1050 | Ferritin | ukb-d-1747_1 | hair color | 122 | 4.799 | 0.049 | 0.039 | 0.212 |
| ieu-a-8 | Coronary heart disease | ukb-d-1747_1 | hair color | 108 | 5.257 | 0.002 | 0.008 | 0.804 |
| ieu-a-1008 | Platelet count | ukb-d-1747_1 | hair color | 127 | 5.121 | 0.000 | 0.001 | 0.866 |
| ieu-a-1000 | Depressive symptoms | ukb-d-1747_1 | hair color | 136 | 5.725 | 0.023 | 0.026 | 0.373 |
| ieu-a-962 | Ever vs never smoked | ukb-d-1747_1 | hair color | 119 | 4.914 | -0.004 | 0.018 | 0.803 |
| ieu-a-1037 | Difference in height between childhood and adulthood | ukb-d-1747_1 | hair color | 116 | 4.951 | 0.008 | 0.018 | 0.637 |
| ieu-a-832 | Rheumatoid arthritis | ukb-d-1747_1 | hair color | 153 | 7.137 | -0.003 | 0.002 | 0.144 |
| ieu-a-1054 | Gout | ukb-d-1747_1 | hair color | 73 | 4.709 | 0.016 | 0.028 | 0.557 |
| ieu-a-1101 | Urinary albumin-to-creatinine ratio | ukb-d-1747_1 | hair color | 108 | 4.551 | 0.106 | 0.078 | 0.171 |
| ieu-a-1183 | ADHD | ukb-d-1747_1 | hair color | 159 | 5.182 | 0.013 | 0.007 | 0.089 |
| ieu-a-67 | Waist circumference | ukb-d-1747_1 | hair color | 168 | 7.804 | 0.015 | 0.014 | 0.256 |
| ieu-a-92 | Obesity class 3 | ukb-d-1747_1 | hair color | 113 | 4.413 | -0.029 | 0.020 | 0.150 |
| ieu-a-996 | Eczema | ukb-d-1747_1 | hair color | 147 | 4.826 | 0.003 | 0.012 | 0.823 |
| ieu-a-298 | Alzheimer's disease | ukb-d-1747_1 | hair color | 420 | 9.326 | 0.001 | 0.002 | 0.601 |
| ieu-a-1171 | Knee osteoarthritis | ukb-d-1747_1 | hair color | 102 | 4.482 | -0.276 | 1.847 | 0.881 |
| ieu-a-836 | College completion | ukb-d-1747_1 | hair color | 126 | 5.758 | -0.001 | 0.010 | 0.905 |
| ieu-a-1045 | Hippocampus volume | ukb-d-1747_1 | hair color | 94 | 4.481 | 0.000 | 0.000 | 0.535 |
| ieu-a-1051 | Transferrin Saturation | ukb-d-1747_1 | hair color | 107 | 5.061 | -0.007 | 0.014 | 0.620 |
| ieu-a-87 | Extreme waist-to-hip ratio | ukb-d-1747_1 | hair color | 56 | 4.571 | 0.004 | 0.056 | 0.942 |
| ieu-a-1009 | Subjective well being | ukb-d-1747_1 | hair color | 129 | 5.786 | 0.032 | 0.020 | 0.117 |
| ieu-a-89 | Height | ukb-d-1747_1 | hair color | 350 | 10.433 | -0.006 | 0.011 | 0.579 |
| ieu-a-1003 | Leptin | ukb-d-1747_1 | hair color | 114 | 4.432 | 0.257 | 0.141 | 0.068 |
| ieu-a-963 | Former vs current smoker | ukb-d-1747_1 | hair color | 96 | 4.863 | -0.011 | 0.011 | 0.310 |
| ieu-a-115 | Neo-extraversion | ukb-d-1747_1 | hair color | 89 | 4.407 | 0.026 | 0.035 | 0.455 |
| ieu-a-29 | Birth length | ukb-d-1747_1 | hair color | 102 | 4.720 | 0.056 | 0.051 | 0.272 |
| ieu-a-804 | Major depressive disorder | ukb-d-1747_1 | hair color | 168 | 4.834 | -0.009 | 0.008 | 0.245 |
| ieu-a-1042 | Nucleus accumbens volume | ukb-d-1747_1 | hair color | 88 | 4.787 | 0.000 | 0.000 | 0.928 |
| ieu-a-117 | Neo-openness to experience | ukb-d-1747_1 | hair color | 102 | 4.678 | 0.003 | 0.012 | 0.814 |
| ieu-a-93 | Overweight | ukb-d-1747_1 | hair color | 112 | 4.924 | -0.011 | 0.016 | 0.482 |
| ieu-a-26 | Type 2 diabetes | ukb-d-1747_1 | hair color | 118 | 5.112 | -0.008 | 0.006 | 0.154 |
| ieu-a-1184 | Autism Spectrum Disorder | ukb-d-1747_1 | hair color | 123 | 4.892 | 0.011 | 0.009 | 0.216 |
| ieu-a-90 | Obesity class 1 | ukb-d-1747_1 | hair color | 94 | 5.395 | -0.004 | 0.008 | 0.578 |
| ieu-a-44 | Asthma | ukb-d-1747_1 | hair color | 107 | 5.174 | 0.007 | 0.005 | 0.141 |
| ieu-a-999 | Body fat | ukb-d-1747_1 | hair color | 113 | 5.894 | -0.021 | 0.009 | 0.020 |
| ieu-a-837 | Years of schooling | ukb-d-1747_1 | hair color | 130 | 5.422 | 0.021 | 0.028 | 0.455 |
| ieu-a-1070 | Sitting height ratio | ukb-d-1747_1 | hair color | 118 | 4.975 | -0.017 | 0.015 | 0.247 |
| ieu-a-987 | Lung cancer | ukb-d-1747_1 | hair color | 168 | 5.583 | -0.001 | 0.006 | 0.876 |
| ieu-a-1006 | Mean platelet volume | ukb-d-1747_1 | hair color | 108 | 7.643 | -0.016 | 0.055 | 0.776 |
| ieu-a-118 | Neuroticism | ukb-d-1747_1 | hair color | 135 | 4.496 | 1.001 | 2.013 | 0.619 |
| ieu-a-113 | Neo-agreeableness | ukb-d-1747_1 | hair color | 89 | 4.582 | 0.040 | 0.055 | 0.469 |
| ieu-a-1048 | Thalamus volume | ukb-d-1747_1 | hair color | 100 | 4.940 | -0.001 | 0.001 | 0.255 |
| ieu-a-116 | Neo-neuroticism | ukb-d-1747_1 | hair color | 71 | 5.104 | 0.002 | 0.001 | 0.160 |
| ieu-a-1189 | Obsessive Compulsive Disorder | ukb-d-1747_1 | hair color | 108 | 4.745 | 0.007 | 0.011 | 0.537 |
| ieu-a-1012 | Plasma cortisol | ukb-d-1747_1 | hair color | 105 | 4.533 | 0.045 | 0.054 | 0.406 |
| ieu-a-1046 | Pallidum volume | ukb-d-1747_1 | hair color | 95 | 4.489 | 0.000 | 0.000 | 0.933 |
| ieu-a-45 | Anorexia nervosa | ukb-d-1747_1 | hair color | 106 | 5.332 | -0.006 | 0.003 | 0.053 |
| ieu-a-988 | Small cell lung carcinoma | ukb-d-1747_1 | hair color | 121 | 4.950 | 0.006 | 0.007 | 0.411 |
| ieu-a-292 | Inflammatory bowel disease | ukb-d-1747_1 | hair color | 422 | 16.914 | 0.000 | 0.001 | 0.970 |
| ieu-a-1043 | Amygdala volume | ukb-d-1747_1 | hair color | 112 | 4.678 | 0.000 | 0.000 | 0.349 |

**Table S5**: Detailed information on the 81 exposures and the outcome used in the negative control study. The causal effect estimates, SEs, and $p$-values obtained using MR-GMM under the threshold $p < 0.05$ are presented.

| Exposure ID | Exposure | Outcome ID | Outcome | Uniprot | Symbol | # of IVs | IV strength | Estimate | SE | P-value |
|---|---|---|---|---|---|---|---|---|---|---|
| prot-a-10 | Histo-blood group ABO system transferase | ieu-a-7 | CHD | P16442 | ABO | 100 | 87.414 | 0.033 | 0.000 | 0.000 |
| prot-a-1074 | Low affinity immunoglobulin gamma Fc region receptor II-a | ieu-a-7 | CHD | P12318 | FCGR2A | 98 | 102.962 | 0.004 | 0.000 | 0.000 |
| prot-a-1075 | Low affinity immunoglobulin gamma Fc region receptor II-b | ieu-a-7 | CHD | P31994 | FCGR2B | 90 | 26.761 | 0.005 | 0.001 | 0.000 |
| prot-a-1112 | Four-jointed box protein 1 | ieu-a-7 | CHD | Q86VR8 | FJX1 | 112 | 4.861 | 0.367 | 0.009 | 0.000 |
| prot-a-1115 | Inactive peptidyl-prolyl cis-trans isomerase FKBP6 | ieu-a-7 | CHD | O75344 | FKBP6 | 112 | 38.743 | 0.024 | 0.000 | 0.000 |
| prot-a-1196 | Growth/differentiation factor 5 | ieu-a-7 | CHD | P43026 | GDF5 | 111 | 16.688 | 0.036 | 0.002 | 0.000 |
| prot-a-1199 | GTP-binding protein GEM | ieu-a-7 | CHD | P55040 | GEM | 109 | 20.534 | 0.034 | 0.000 | 0.000 |
| prot-a-1277 | Granulins | ieu-a-7 | CHD | P28799 | GRN | 87 | 15.172 | 0.119 | 0.000 | 0.000 |
| prot-a-1397 | Intercellular adhesion molecule 1 | ieu-a-7 | CHD | P05362 | ICAM1 | 122 | 68.277 | -0.003 | 0.001 | 0.000 |
| prot-a-1400 | Intercellular adhesion molecule 4 | ieu-a-7 | CHD | Q14773 | ICAM4 | 106 | 4.511 | -0.463 | 0.142 | 0.001 |
| prot-a-1502 | Interleukin-1 receptor-like 1 | ieu-a-7 | CHD | Q01638 | IL1RL1 | 115 | 20.308 | 0.017 | 0.001 | 0.000 |
| prot-a-1540 | Interleukin-6 receptor subunit alpha | ieu-a-7 | CHD | P08887 | IL6R | 105 | 52.887 | -0.044 | 0.000 | 0.000 |
| prot-a-1741 | Leukocyte immunoglobulin-like receptor subfamily A member 6 | ieu-a-7 | CHD | Q6PI73 | LILRA6 | 90 | 4.428 | -0.404 | 0.127 | 0.001 |
| prot-a-18 | Low molecular weight phosphotyrosine protein phosphatase | ieu-a-7 | CHD | P24666 | ACP1 | 89 | 59.839 | -0.017 | 0.000 | 0.000 |
| prot-a-180 | Arf-GAP with SH3 domain, ANK repeat and PH domain-containing protein 2 | ieu-a-7 | CHD | O43150 | ASAP2 | 100 | 10.746 | 0.052 | 0.003 | 0.000 |
| prot-a-1863 | Mannose-binding protein C | ieu-a-7 | CHD | P11226 | MBL2 | 114 | 19.979 | -0.022 | 0.001 | 0.000 |
| prot-a-1902 | MICOS complex subunit MIC10 | ieu-a-7 | CHD | Q5TGZ0 | MICOS10 | 97 | 73.117 | 0.021 | 0.000 | 0.000 |
| prot-a-1949 | Beta-microseminoprotein | ieu-a-7 | CHD | P08118 | MSMB | 119 | 17.924 | 0.030 | 0.003 | 0.000 |
| prot-a-2023 | NADH dehydrogenase [ubiquinone] 1 beta subcomplex subunit 4 | ieu-a-7 | CHD | O95168 | NDUFB4 | 106 | 11.173 | 0.044 | 0.005 | 0.000 |
| prot-a-2025 | NADH dehydrogenase [ubiquinone] iron-sulfur protein 4, mitochondrial | ieu-a-7 | CHD | O43181 | NDUFS4 | 94 | 12.264 | -0.033 | 0.003 | 0.000 |
| prot-a-2095 | Neuregulin-1 | ieu-a-7 | CHD | Q02297 | NRG1 | 95 | 4.686 | -0.316 | 0.069 | 0.000 |
| prot-a-2230 | Platelet-derived growth factor receptor beta | ieu-a-7 | CHD | P09619 | PDGFRB | 88 | 26.856 | -0.004 | 0.000 | 0.000 |
| prot-a-2247 | Proenkephalin-A | ieu-a-7 | CHD | P01210 | PENK | 105 | 8.102 | -0.023 | 0.003 | 0.000 |
| prot-a-2284 | Group XIIB secretory phospholipase A2-like protein | ieu-a-7 | CHD | Q9BX93 | PLA2G12B | 97 | 6.633 | 0.222 | 0.008 | 0.000 |
| prot-a-2291 | Secretory phospholipase A2 receptor | ieu-a-7 | CHD | Q13018 | PLA2R1 | 101 | 27.242 | 0.002 | 0.000 | 0.000 |
| prot-a-2499 | RNA binding protein fox-1 homolog 2 | ieu-a-7 | CHD | O43251 | RBFOX2 | 80 | 4.433 | 0.624 | 0.176 | 0.000 |
| prot-a-2575 | rRNA methyltransferase 3, mitochondrial | ieu-a-7 | CHD | Q9HC36 | MRM3 | 98 | 11.949 | 0.046 | 0.000 | 0.000 |
| prot-a-2731 | Sialic acid-binding Ig-like lectin 9 | ieu-a-7 | CHD | Q9Y336 | SIGLEC9 | 88 | 116.985 | -0.009 | 0.000 | 0.000 |
| prot-a-2732 | Tyrosine-protein phosphatase non-receptor type substrate 1 | ieu-a-7 | CHD | P78324 | SIRPA | 94 | 70.662 | -0.003 | 0.000 | 0.000 |
| prot-a-276 | ADP-ribosyl cyclase/cyclic ADP-ribose hydrolase 2 | ieu-a-7 | CHD | Q10588 | BST1 | 101 | 9.825 | -0.029 | 0.002 | 0.000 |
| prot-a-2829 | Spondin-1 | ieu-a-7 | CHD | Q9HCB6 | SPON1 | 108 | 6.638 | 0.032 | 0.009 | 0.001 |
| prot-a-2940 | Teratocarcinoma-derived growth factor 1 | ieu-a-7 | CHD | P13385 | CRIPTO | 106 | 14.723 | 0.015 | 0.000 | 0.000 |
| prot-a-2941 | Tyrosyl-DNA phosphodiesterase 1 | ieu-a-7 | CHD | Q9NUW8 | TDP1 | 95 | 4.340 | 0.429 | 0.126 | 0.001 |
| prot-a-303 | Complement C1q tumor necrosis factor-related protein 1 | ieu-a-7 | CHD | Q9BXJ1 | C1QTNF1 | 98 | 9.776 | 0.153 | 0.001 | 0.000 |
| prot-a-3214 | Vitronectin | ieu-a-7 | CHD | P04004 | VTN | 101 | 39.214 | 0.025 | 0.000 | 0.000 |
| prot-a-426 | CD209 antigen | ieu-a-7 | CHD | Q9NNX6 | CD209 | 97 | 9.276 | 0.054 | 0.005 | 0.000 |
| prot-a-540 | Chitinase-3-like protein 1 | ieu-a-7 | CHD | P36222 | CHI3L1 | 96 | 39.579 | 0.014 | 0.000 | 0.000 |
| prot-a-570 | C-type lectin domain family 12 member A | ieu-a-7 | CHD | Q5QGZ9 | CLEC12A | 108 | 88.240 | -0.001 | 0.000 | 0.000 |
| prot-a-635 | COP9 signalosome complex subunit 2 | ieu-a-7 | CHD | P61201 | COPS2 | 108 | 11.106 | 0.050 | 0.005 | 0.000 |
| prot-a-643 | Carboxypeptidase A4 | ieu-a-7 | CHD | Q9UI42 | CPA4 | 99 | 60.600 | 0.013 | 0.000 | 0.000 |
| prot-a-854 | Dedicator of cytokinesis protein 9 | ieu-a-7 | CHD | Q9BZ29 | DOCK9 | 124 | 4.695 | 0.391 | 0.106 | 0.000 |
| prot-a-91 | Anaphase-promoting complex subunit 7 | ieu-a-7 | CHD | Q9UJX3 | ANAPC7 | 89 | 13.040 | -0.036 | 0.004 | 0.000 |
| prot-a-96 | Angiopoietin-related protein 1 | ieu-a-7 | CHD | O95841 | ANGPTL1 | 103 | 7.317 | -0.064 | 0.004 | 0.000 |
| prot-a-971 | Endoplasmic reticulum aminopeptidase 1 | ieu-a-7 | CHD | Q9NZ08 | ERAP1 | 102 | 22.514 | -0.023 | 0.003 | 0.000 |
| prot-a-972 | Endoplasmic reticulum aminopeptidase 2 | ieu-a-7 | CHD | Q6P179 | ERAP2 | 104 | 42.251 | 0.023 | 0.000 | 0.000 |

**Table S6**: The identified 45 CHD-associated plasma proteins. The proteins are annotated with Uniprot accession and gene symbols. The causal effect estimates, SEs, and $p$-values obtained using MR-GMM under the threshold $p < 0.05$ are presented.
